# Implementing long-acting injectable HIV pre-exposure prophylaxis services at private pharmacies in Kenya: Client, pharmacy provider, and key stakeholder perspectives on potential challenges and opportunities

**DOI:** 10.1101/2025.02.03.25321360

**Authors:** Stephanie Roche, Kevin Kamolloh, Nicholas Thuo, Maurice Opiyo, Vallery Ogello, Alfred Odira, Emmah Owidi, Perez Ochwal, Marion Hewa, Lydia Adiema, Felix Mogaka, Victor O. Omollo, Rachel C. Malen, Kendall Harkey, Jenell Stewart, Elizabeth A. Bukusi, Kenneth Ngure, Katrina F. Ortblad

## Abstract

**Introduction:** Maximizing the impact of new and forthcoming long-acting injectable HIV pre-exposure prophylaxis (PrEP) products will require novel delivery approaches that widen accessibility and prioritize clients’ needs and preferences. To understand the potential barriers and facilitators to delivering injectable PrEP via private pharmacies in Kenya, we conducted qualitative formative research.

**Methods:** From July to September 2023, we interviewed pharmacy providers, pharmacy clients, and other key stakeholders of HIV service delivery in Central and Western Kenya. Our purposive sample included some providers and clients with prior experience delivering or obtaining oral PrEP at a pharmacy as part of a pilot study and some without such experience. We analyzed verbatim transcripts thematically using a combination of inductive and deductive approaches, the latter informed by the Consolidated Framework for Implementation Research (CFIR).

**Results:** We interviewed 25 pharmacy clients, 16 pharmacy providers, and 9 key stakeholders. Each group was ∼50% female, and median age among clients was 25 (IQR 23-29). Overall, participants supported the idea of pharmacy-based injectable PrEP delivery. Anticipated facilitators included perceived benefits of injectable over oral PrEP; characteristics of pharmacies (e.g., long operating hours) that could fulfill clients’ need for accessible, fast, and private injectable PrEP services; and existing skillsets of pharmacy providers, especially those already trained on injectable contraception. Anticipated barriers included gaps in enabling policy; pharmacies’ lack of integration with the public health sector, such as its health information system; low client knowledge of and/or ability to pay for injectable PrEP; and pharmacy staffing and compensation structures that could disincentivize providers.

**Conclusions:** Participants in this study expressed cautious optimism that private pharmacies could be an effective delivery channel for injectable PrEP in Kenya. If private pharmacies facilitate access to and use of injectable PrEP, they could play a pivotal role in ending HIV as a public health threat.

## INTRODUCTION

After a decade of suboptimal access and uptake of oral HIV pre-exposure prophylaxis (PrEP) worldwide,^1^ the advent of highly effective long-acting injectable PrEP products has brought renewed optimism to the HIV prevention landscape and a keen interest in novel delivery approaches that might accommodate multiple PrEP products while supporting clients’ informed choice.^2, 3^ To date, 53 countries—including 13 in sub-Saharan Africa (SSA)—have approved bimonthly intramuscular injections of long-acting cabotegravir (CAB-LA) for HIV prevention,^4^ and recent evidence from the PURPOSE 1 and 2 trials suggest that twice-yearly subcutaneous lenacapavir injections may become part of the HIV prevention toolkit in the near future.^5, 6^ These products may be especially impactful in SSA, where half of the world’s new HIV infections (∼660,000/1.3 million) occurred in 2022.^7^

To accelerate access to injectable PrEP, HIV prevention advocates have called for its roll-out to incorporate learnings from that of oral PrEP.^4^ For example, to accommodate a known preference among many clients to access PrEP in non-clinical settings,^8^ countries will need to adapt where injectable PrEP services are delivered and by whom, in line with principles of differentiated service delivery.^9, 10^ Another key lesson learned from oral PrEP is that limiting initial roll-out to key populations (e.g., sex workers) can create PrEP stigma.^11, 12^ To avoid this, some have called for countries with generalized HIV epidemics to “normalize” injectable PrEP^13^ by availing it from the outset to members of general population who could benefit—as Zambia^14^ is doing—and delivering it in places where the general population accesses health services.

Due to global shortages of CAB-LA,^14^ its programmatic roll-out is still nascent. Currently, most countries with CAB-LA supply are prioritizing traditional clinical settings for initial roll-out, with injections administered by nurses or doctors. ^15–17^ However, early examples of differentiated CAB-LA delivery in the United States include injection administration task-shifted to community health workers at a single primary care center in Washington, D.C.^18^ and to pharmacists at a single pharmacy in Seattle, Washington.^19^

Faced with declining donor funding for its national HIV program,^20, 21^ Kenya is exploring ways to leverage the private sector for HIV service delivery,^22^ including the largely untapped well of human and physical resources comprising its private pharmacy sector.^22, 23^ This sector boasts approximately 11,000 licensed pharmaceutical technologists; 2,600 licensed pharmacists; and 7,400 registered pharmacies^24^ routinely accessed by both the general and key populations for sexual and reproductive health products, such as condoms and emergency contraception. Moreover, recent pilot studies of pharmacy-delivered oral PrEP services in Kenya have observed high uptake and acceptability among clients and providers.^25–28^ With approval and programmatic roll-out of injectable PrEP in Kenya on the horizon, understanding the potential barriers and facilitators to its delivery via private pharmacies is of timely relevance.

## METHODS

### Setting and participants

This study took place in Kisumu and Kiambu Counties of Kenya where HIV prevalence is 15% and 2%, respectively.^29^ Informed by the 7Ps Framework,^30^ we used quota sampling to purposively select PrEP clients, pharmacy providers, and other key stakeholders (all age 18+). Eligible clients self-reported purchasing health products at pharmacies and using oral PrEP within the past six months. Eligible pharmacy providers were licensed pharmacists or pharmaceutical technologists (hereafter, “pharm techs”). Eligible stakeholders were individuals with direct involvement in HIV- or pharmacy-related policymaking, regulation, and/or program implementation in Kenya.

To capture perspectives from individuals with prior experience obtaining or delivering oral PrEP services at pharmacies, we drew two-thirds of clients and half of providers from individuals who had participated in a pilot study^26^ six months prior; research assistants (RAs) randomly selected providers as well as clients who refilled and never refilled PrEP during the pilot, and contacted them via phone. The remaining PrEP clients and pharmacy providers were recruited in person from public health facilities and licensed pharmacies in the study counties. For our client sample, we aimed to have an even distribution of men and women above and below 25 years old. Stakeholders were recruited through the research team’s professional networks via phone or email. Given the tightly scoped nature of our research question, we anticipated achieving information power^31^ with 8 to 10 interviews per subcategory, for a total of ∼50 interviews.

### Data collection

We developed semi-structured interview guides (**Appendix A**) informed by the updated Consolidated Framework for Implementation Research (CFIR)^32^ and findings from previous studies on pharmacy-delivered oral PrEP services in Kenya.^25, 26, 33, 34^ The guides included a brief, standardized description of CAB-LA and lenacapavir (e.g., injection type and site, common side effects) and asked about anticipated willingness to obtain, deliver, or support injectable PrEP services at pharmacies, potential barriers, and possible solutions or recommendations. Experienced Kenyan qualitative RAs conducted interviews in participants’ preferred language (English, Swahili, or Dholuo) in a private location of the participant’s choosing or over the phone. Interviews were audio recorded and transcribed verbatim. After each interview, RAs wrote debrief reports summarizing key points.

### Analysis

We analyzed data thematically using a combination of inductive and deductive approaches, informed by principles of rapid qualitative analysis.^35, 36^ After repeated readings of debrief reports, author SDR drafted a memo template organized around CFIR constructs, which was refined based on RA feedback. Authors NT, MO, VO, AO, PO, MH, and LA developed one memo per interview comprised of a summary, illustrative quotes, and observations about early patterns. Memos were entered into an electronic data collection platform (REDCap) and exported in matrix format. Authors SDR, KH, and RM read across and within memos independently to identify additional patterns; consulted original transcripts for context, as needed; and collapsed or expanded key categories through consensus.

### Patient and public involvement and ethics

Neither patients nor the public were directly involved in the design or conduct of this study; however, the focus of this research—understanding the potential barriers and facilitators to implementing injectable PrEP delivery at private pharmacies—is responsive to broader calls from patients and HIV advocacy groups to make PrEP services available outside of traditional clinic settings. Our study protocol was approved by the institutional review boards of the Kenya Medical Research Institute and University of Washington. All participants completed written informed consent in English, Kiswahili, or Dholuo, and were compensated 500 Kenyan shillings (∼$5 US dollars [USD]) per interview.

## RESULTS

### Participants

From July to September 2023, we interviewed 24 PrEP clients, 16 pharmacy providers, and nine key stakeholders (**Table 1**). About half of clients were female and under the age of 25. Among clients who never refilled PrEP at the pharmacy, all attributed their discontinuation to personal circumstances (e.g., change in perceived HIV risk) and not to the delivery venue. About two-thirds of pharmacy providers were female, and their median age was 35 years. Among stakeholders, two-thirds were female, median age was 44, and about half worked for government and half for non-governmental organizations.

**Table 1.** Demographics of participants who completed in-depth interviews.

| Characteristic | PrEP clients<br>(N=24) | Pharmacy providers<br>(N=16) | Other key stakeholders<br>(N=9) |
| --- | --- | --- | --- |
| Female<sup>a</sup> | 13 (54%) | 10 (63%) | 6 (67%) |
| Age<sup>b</sup>, median (IQR) | 25 (23-29) | 35 (33-38) | 44 (38-51) |
| Unmarried | 17 (71%) |  |  |
| Education: Secondary or higher | 20 (83%) | 16 (100%) |  |
| Prior oral PrEP experience<sup>a</sup> |  |  |  |
| *Clinic-delivered services only* | 8 (33%) |  |  |
| *Pharmacy-delivered services, never refilled<sup>c,d</sup>* | 8 (33%) |  |  |
| *Pharmacy-delivered services, refilled 1+ time<sup>c</sup>* | 8 (33%) |  |  |
| Cadre |  |  |  |
| *Pharmaceutical technologist* |  | 11 (69%) |  |
| *Pharmacist* |  | 5 (31%) |  |
| Prior experience delivering oral PrEP<sup>a,c</sup> |  | 8 (50%) |  |
| Years in pharmacy work, median (IQR) |  | 11 (7-13) |  |
| Current or prior experience administering injections |  | 13 (81%) |  |
| Their pharmacy sells and/or administers the following injection(s): |  |  |  |
| *Injectable contraception* |  | 12 (75%) |  |
| *Tetanus vaccine* |  | 7 (44%) |  |
| *Rabies vaccine* |  | 7 (44%) |  |
| *Other<sup>e</sup>* |  | 5 (31%) |  |
| Employer |  |  |  |
| *Kenya Ministry of Health* |  |  | 3 (33%) |
| *Pharmacy professional body* |  |  | 3 (33%) |
| *Pharmacy regulator* |  |  | 2 (22%) |
| *PrEP implementing partner* |  |  | 1 (11%) |

<sup>a</sup> Characteristic used during purposive sampling to achieve the distribution shown.
<sup>b</sup> Client interviewees purposively sampled so half were under 25 years old.
<sup>c</sup> As part of the Pharm PrEP Pilot Extension Study, which ran from January-July 2022 at 12 pharmacies in Kisumu and Kiambu Counties, Kenya <sup>d</sup>Among these 8 clients, self-reported reasons for not refilling oral PrEP at the pharmacy included: reduced HIV risk so felt PrEP no longer needed (n=2); moved to new county & travel distance too far (n=2); inadvertently missed follow-up appointment (n=2; of note, these clients only managed to return to the pharmacy after the study had ended and were referred to nearby public health facilities, where both re-initiated PrEP services); pill burden (n=1); and PrEP side effects (n=1).
<sup>e</sup> Other injections offered at these five providers' pharmacies include anti-D immunoglobulin injection, insulin injection, injectable painkillers (e.g., tramadol), injectable antibiotics (e.g., metronidazole), and vaccines for flu and hepatitis.

Below, we present the potential barriers and facilitators participants identified, with those pertaining to the CFIR domains of Innovation, Outer Setting, and Inner Setting summarized in **Table 2** and those pertaining to the Individuals domain summarized in **Table 3**.

**Table 2.**
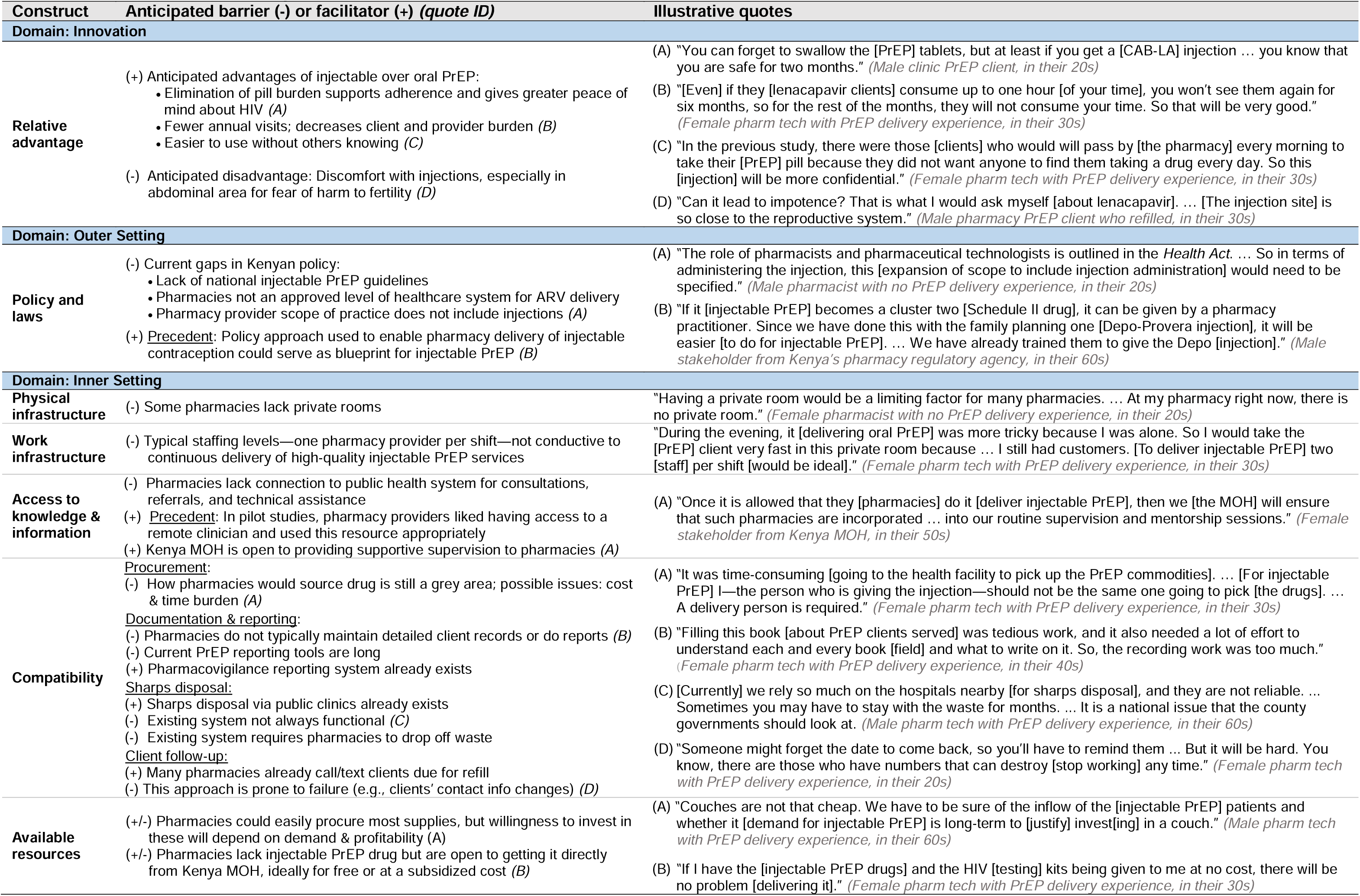
Potential barriers and facilitators to pharmacy-delivered injectable PrEP in the innovation, outer setting, and inner setting domains of the Consolidated Framework for Implementation Research.

| Construct | Anticipated barrier (-) or facilitator (+) (quote ID) | Illustrative quotes |
| --- | --- | --- |
| **Domain: Innovation** |  |  |
| **Relative advantage** | (+) Anticipated advantages of injectable over oral PrEP:<br>• Elimination of pill burden supports adherence and gives greater peace of mind about HIV (A)<br>• Fewer annual visits; decreases client and provider burden (B)<br>• Easier to use without others knowing (C)<br>(-) Anticipated disadvantage: Discomfort with injections, especially in abdominal area for fear of harm to fertility (D) | (A) “You can forget to swallow the [PrEP] tablets, but at least if you get a [CAB-LA] injection ... you know that you are safe for two months.” (*Male clinic PrEP client, in their 20s*)<br>(B) “[Even] if they [lenacapavir clients] consume up to one hour [of your time], you won’t see them again for six months, so for the rest of the months, they will not consume your time. So that will be very good.” (*Female pharm tech with PrEP delivery experience, in their 30s*)<br>(C) “In the previous study, there were those [clients] who would will pass by [the pharmacy] every morning to take their [PrEP] pill because they did not want anyone to find them taking a drug every day. So this [injection] will be more confidential.” (*Female pharm tech with PrEP delivery experience, in their 30s*)<br>(D) “Can it lead to impotence? That is what I would ask myself [about lenacapavir]. ... [The injection site] is so close to the reproductive system.” (*Male pharmacy PrEP client who refilled, in their 30s*) |
| **Domain: Outer Setting** |  |  |
| **Policy and laws** | (-) Current gaps in Kenyan policy:<br>• Lack of national injectable PrEP guidelines<br>• Pharmacies not an approved level of healthcare system for ARV delivery<br>• Pharmacy provider scope of practice does not include injections (A)<br>(+) <u>Precedent</u>: Policy approach used to enable pharmacy delivery of injectable contraception could serve as blueprint for injectable PrEP (B) | (A) “The role of pharmacists and pharmaceutical technologists is outlined in the *Health Act*. ... So in terms of administering the injection, this [expansion of scope to include injection administration] would need to be specified.” (*Male pharmacist with no PrEP delivery experience, in their 20s*)<br>(B) “If it [injectable PrEP] becomes a cluster two [Schedule II drug], it can be given by a pharmacy practitioner. Since we have done this with the family planning one [Depo-Provera injection], it will be easier [to do for injectable PrEP]. ... We have already trained them to give the Depo [injection].” (*Male stakeholder from Kenya’s pharmacy regulatory agency, in their 60s*) |
| **Domain: Inner Setting** |  |  |
| **Physical infrastructure** | (-) Some pharmacies lack private rooms | “Having a private room would be a limiting factor for many pharmacies. ... At my pharmacy right now, there is no private room.” (*Female pharmacist with no PrEP delivery experience, in their 20s*) |
| **Work infrastructure** | (-) Typical staffing levels—one pharmacy provider per shift—not conducive to continuous delivery of high-quality injectable PrEP services | “During the evening, it [delivering oral PrEP] was more tricky because I was alone. So I would take the [PrEP] client very fast in this private room because ... I still had customers. [To deliver injectable PrEP] two [staff] per shift [would be ideal].” (*Female pharm tech with PrEP delivery experience, in their 30s*) |
| **Access to knowledge & information** | (-) Pharmacies lack connection to public health system for consultations, referrals, and technical assistance<br>(+) <u>Precedent</u>: In pilot studies, pharmacy providers liked having access to a remote clinician and used this resource appropriately<br>(+) Kenya MOH is open to providing supportive supervision to pharmacies (A) | (A) “Once it is allowed that they [pharmacies] do it [deliver injectable PrEP], then we [the MOH] will ensure that such pharmacies are incorporated ... into our routine supervision and mentorship sessions.” (*Female stakeholder from Kenya MOH, in their 50s*) |
| **Compatibility** | <u>Procurement</u>:<br>(-) How pharmacies would source drug is still a grey area; possible issues: cost & time burden (A)<br><u>Documentation & reporting</u>:<br>(-) Pharmacies do not typically maintain detailed client records or do reports (B)<br>(-) Current PrEP reporting tools are long<br>(+) Pharmacovigilance reporting system already exists<br><u>Sharps disposal</u>:<br>(+) Sharps disposal via public clinics already exists<br>(-) Existing system not always functional (C)<br>(-) Existing system requires pharmacies to drop off waste<br><u>Client follow-up</u>:<br>(+) Many pharmacies already call/text clients due for refill<br>(-) This approach is prone to failure (e.g., clients’ contact info changes) (D) | (A) “It was time-consuming [going to the health facility to pick up the PrEP commodities]. ... [For injectable PrEP] I—the person who is giving the injection—should not be the same one going to pick [the drugs]. ... A delivery person is required.” (*Female pharm tech with PrEP delivery experience, in their 30s*)<br>(B) “Filling this book [about PrEP clients served] was tedious work, and it also needed a lot of effort to understand each and every book [field] and what to write on it. So, the recording work was too much.” (*Female pharm tech with PrEP delivery experience, in their 40s*)<br>(C) [Currently] we rely so much on the hospitals nearby [for sharps disposal], and they are not reliable. ... Sometimes you may have to stay with the waste for months. ... It is a national issue that the county governments should look at. (*Male pharm tech with PrEP delivery experience, in their 60s*)<br>(D) “Someone might forget the date to come back, so you’ll have to remind them ... But it will be hard. You know, there are those who have numbers that can destroy [stop working] any time.” (*Female pharm tech with PrEP delivery experience, in their 20s*) |
| **Available resources** | (+/-) Pharmacies could easily procure most supplies, but willingness to invest in these will depend on demand & profitability (A)<br>(+/-) Pharmacies lack injectable PrEP drug but are open to getting it directly from Kenya MOH, ideally for free or at a subsidized cost (B) | (A) “Couches are not that cheap. We have to be sure of the inflow of the [injectable PrEP] patients and whether it [demand for injectable PrEP] is long-term to [justify] invest[ing] in a couch.” (*Male pharm tech with PrEP delivery experience, in their 60s*)<br>(B) “If I have the [injectable PrEP drugs] and the HIV [testing] kits being given to me at no cost, there will be no problem [delivering it].” (*Female pharm tech with PrEP delivery experience, in their 30s*) |

**Table 3.** Potential barriers and facilitators to pharmacy-delivered injectable PrEP in the individuals domain of the Consolidated Framework for Implementation Research.

| Construct | Anticipated barrier (-) or facilitator (+) (quote ID) | Illustrative quotes |
| --- | --- | --- |
| Client need | (+) Pharmacy delivery would fulfill some clients' need for HIV services that are:<br>• Accessible (A)<br>• Fast/efficient (B)<br>• Private/discreet<br>• Respectful/nonjudgmental | (A) "It is easier [getting PrEP at a pharmacy]. Most pharmacies close around 8 or 9pm ... and are open throughout the week, unlike clinics, which are open maybe 5 days a week ... and maybe you go to get [PrEP] and you find that they have closed already." (*Male pharmacy PrEP client who refilled, in their 40s*)<br>(B) "[At public clinics] you will have to go through three people before you reach the doctor who is prescribing the medication to you. But if you go to the pharmacy, you go directly to the person who will serve you what you want." (*Male pharmacy PrEP client who refilled, in their 20s*) |
| Client capability | (-) Low client awareness of injectable PrEP<br>(-) Low client awareness of its availability at pharmacies (A) | (A) "People won't know such [injectable PrEP] is offered at the pharmacies. They only know of the hospitals. ... So we need a bit of advertising or maybe a banner." (*Male pharmacist with prior PrEP delivery experience, in their 30s*) |
| Client opportunity | (+/-) Cost: Amount pharmacies charge will influence affordability for clients; government could subsidize delivery and put price caps (A)<br>(+/-) Quantity & geographic distribution of pharmacies offering injectable PrEP will influence clients' access (B) | (A) "You can never tell how much a chemist will ask for. ... They might say they will be charging 200 [shillings], and you might fail to get that 200. So it will be a problem for those of us who don't have the ability to pay." (*Female pharmacy PrEP client who refilled, in their 30s*)<br>(B) "When I came to [the pharmacy to] pick those [oral PrEP] medications, it was when I had activities to do in [this area]. It needed transport because it is a bit far. ... These PrEPs should be available in many other pharmacies. ... Every region should have these injectables. They should not just be available in one place." (*Male pharmacy PrEP client who refilled, in their 20s*) |
| Client motivation | (+/-) Trust in injectable PrEP (A)<br>(+/-) Confidence in provider's integrity (B)<br>(+/-) Confidence in provider's competence, especially knowledge, counseling, and injection skills (C) | (A) "[I'd need to know] if it prevents [HIV] 100% and if it's safe." (*Female pharmacy PrEP client who refilled, in their 20s*)<br>(B) "[Providers] should not be someone who sees me in town and starts saying, 'That is my client. I gave that one medicine.' ... I would never go there." (*Male clinic PrEP client, in their 20s*)<br>(C) [The provider] should be able to tell you the benefits of injectable PrEP over oral pills. They must explain to you why they have upgraded from the oral pills to this [injectable] one. (*Female pharmacy PrEP client who did not refill, in their 20s*) |
| Provider capability | (+) For *lenacapavir*: Some pharmacy providers already administer subcutaneous injections; a few have been trained by the MOH to give injectable contraception (A)<br>(+/-) A small number of pharmacies have nurses/doctors; requiring these cadres would disqualify most pharmacies (B)<br>(+) Existing trainings (e.g., on injectable contraception delivery) could be repurposed | (A) "Most licensed pharmacies already offer subcutaneous injections, like insulin. So the knowledge is there. What we are lacking is the [lenacapavir] drug." (*Male pharmacist with no PrEP delivery experience, in their 30s*)<br>(B) "You can have a nurse [administer the injection]. ... I have two licensed nurses [on staff], but I think not all pharmacies can afford to do what we are doing." (*Male pharm tech with PrEP delivery experience, in their 30s*) |
| Provider opportunity | (-) Amount of time it takes to deliver injectable PrEP may, at times, exceed what pharmacy providers have available | "Because of the high cost of overheads, we want to serve as many people in a day as possible and also to reduce the time of serving patients. ... If you're to go through the whole process of having to test patients for HIV and to counsel them on the injectable and everything else, it means taking your time with one patient, [which] I think would be a challenge." (*Male pharmacist with no PrEP delivery experience, in their 20s*) |
| Provider motivation | (-) Low profit margin and high time burden could weaken pharmacy commitment to delivery (A)<br>(-/+) A commission-based compensation structure might disincentivize providers from delivering injectable PrEP; an incentive paid directly to the provider delivering the service might counteract this issue (B) | (A) "A pharmacy is a business. I can't spend time with you as a client for, like, 1 hour when I could have made maybe 10,000 or 5,000 shillings selling [other] medicine." (*Female pharm tech with PrEP delivery experience, in their 30s*)<br>(B) "We work with these [sales] targets. If you achieve your target, there is a certain amount of money [bonus commission] you are given ... So if [after spending time delivering injectable PrEP] there is something you are compensated [directly] at the end of it all yourself, ... that will give me more comfort. Even if I don't get the other one [the bonus commission], I still get something out of this [injectable PrEP delivery]." (*Female pharm tech with PrEP delivery experience, in their 40s*) |

### Innovation domain

#### Relative advantage

Participants anticipated three key advantages of injectable PrEP compared to oral PrEP. First, they thought the elimination of daily pill-taking would facilitate adherence and bring clients greater peace of mind about acquiring HIV. Second, they thought that twice-yearly, compared to quarterly, clinic visits would reduce burden on clients and providers, making it easier for them to access or deliver services. Lastly, they felt injections would be more discrete than oral pills, making it easier for clients to keep their PrEP use private.

However, several participants anticipated that some clients would continue to prefer oral PrEP over CAB-LA due to discomfort exposing part of their buttocks in a pharmacy setting. For lenacapavir, nearly all clients said they would be hesitant to get an injection in the abdominal area due to fear of adverse effects on fertility:

> [The abdomen is] the same place where I carry my pregnancy. If at all I could have been pregnant and still not aware, and they inject that place, I am afraid it may affect me. The one for the buttocks [CAB-LA] is better. *(Female clinic PrEP client, in their 20s)*

### Outer setting domain

#### Policy and laws

Participants identified gaps in Kenyan policy that would need to be addressed to enable pharmacy delivery of injectable PrEP. They called for developing injectable PrEP guidelines that specify pharmacies as a target delivery channel and which cadres should deliver it, noting that for pharmacy providers to deliver it would likely require expanding pharmacists’ and/or pharm techs’ scope of practice to include injection administration. A few stakeholders suggested using an approach similar to that which enabled pharmacy-delivered injectable contraception in Kenya—rescheduling the drug so it can be initiated by pharmacy providers and creating a certificate training program. Participants also reported that Kenya’s current essential medicine list would need to be revised to include injectable PrEP and indicate that these drugs should be available for use (i.e., distributed, stored, prescribed, and dispensed) at the pharmacy level of the healthcare system. Lastly, participants reported the need to establish minimum criteria that pharmacies must meet to deliver injectable PrEP and recommended these include a current government license and registration, private consultation room, and a provider certified in injectable PrEP delivery.

### Inner setting domain

#### Physical infrastructure

Nearly all participants said that pharmacies should be required to have a fully private room to ensure client privacy and comfort, but noted that this requirement might disqualify a considerable number of pharmacies in Kenya.

#### Work infrastructure

Participants reported that most pharmacies have only one pharmacy provider working per shift and identified this staffing level as a potential barrier to making injectable PrEP services available continuously throughout the day.

#### Access to knowledge and information

Nearly all stakeholders and providers—especially those with prior experience delivering oral PrEP—emphasized that pharmacy providers should have access to a clinician with PrEP expertise for on-demand consultations and referrals. Some reported that routine technical assistance and continuing medical education on injectable PrEP delivery would be key to safeguarding quality of care and recommended that training updates carry continuing professional development points, which pharmacy providers need to accrue annually for re-licensure.

### Compatibility

Participants identified four grey areas that could influence how well injectable PrEP delivery fits with how pharmacies typically operate.

#### Procurement

Participants noted the need to establish how pharmacies would procure injectable PrEP and at what price, anticipating that high cost and an overly burdensome procurement process could hinder pharmacies’ uptake. Contrary to how pharmacies in Kenya usually source their drugs— from a wide variety of distributors and manufacturers—some stakeholders recommended pharmacies be required to source injectable PrEP from the government or a limited set of authorized distributors to control drug quality and reduce the risk of stock-outs. Referencing the procurement arrangement used in prior pilot studies^25, 26^—whereby pharmacies retrieved free oral PrEP from nearby public health facilities—several providers welcomed the idea of getting free injectable PrEP from government stock, but called for it to be delivered directly to their pharmacy, in line with how other products reach them.

#### Documentation and reporting

Stakeholders emphasized the importance of pharmacies documenting and reporting injectable PrEP services rendered both for monitoring and evaluation purposes and supply chain forecasting and quantification; however, several pointed out that maintaining detailed client records and completing routine MOH reports would be new for most pharmacies. Noting that pharmacies are not currently integrated into Kenya’s health information system, these participants highlighted the need to establish what metrics pharmacies would report on and how. A few anticipated that the time-consuming nature of these tasks would not fit well within pharmacy workflows, with one stakeholder recommending that existing reporting tools be pared down for pharmacies. Anticipating data quality issues, some stakeholders called for routine auditing of pharmacies’ injectable PrEP data and, where needed, supportive supervision. For pharmacovigilance reporting, providers and stakeholders felt that the existing electronic reporting system managed by the country’s drug regulatory authority would suffice.

#### Sharps disposal

Several providers felt that their pharmacy’s current way of disposing used sharps—bringing them to nearby public health facilities—would work well for injectable PrEP delivery; however, a few anticipated challenges—such as facilities not accepting the waste and the time burden associated with this task—and called for the MOH to ensure the existing sharps disposal system is reliable and to add a pick-up service.

#### Client follow-up

When asked about appointment reminders for injectable PrEP clients, most providers felt their current practices—most commonly, noting the client’s next appointment date in their internal records and calling or texting those who miss their appointment—would suffice; however, some noted that this follow-up approach relies on having up-to-date contact information for the client yet clients often change phone numbers without notifying the pharmacy. Recommendations to mitigate this barrier included issuing appointment cards and asking clients to provide additional contact information, such as their home address, email, or the phone number of a trusted friend or next of kin. One provider recommended pharmacies be equipped with an automated SMS appointment reminder system. One stakeholder similarly called for automated appointment reminders and additionally recommended collaboration with community health workers:

> Human beings are prone to forgetting. So we don’t want somebody writing on a book that, “I need to remember to call this person after six months.” There’s a high likelihood that they will not remember. So how do we leverage on digital systems to support with [appointment] reminders? … My recommendation would be for the pharmacy to invest in good follow-up systems. … [It could involve] phone calls and figuring out how to link to existing community systems. Like the community health promoters—if they can get the unit [address] where the person is staying, … [they could] do [at-home] follow-up just to find out whether the client is still at risk or are they covered using any other HIV prevention methods. *(Female stakeholder from implementing organization, in their 30s)*

### Available resources

When discussing the supplies and equipment they anticipated needing to deliver injectable PrEP (**Table 4**), providers reported that most items would be easy to acquire and/or keep in stock, but several noted that pharmacies’ willingness and ability to invest in these items would depend on the demand for and profitability of injectable PrEP. A few providers and stakeholders recommended the government support pharmacies with the requisite commodities (including the injectable PrEP drug) for free or at a subsidized cost to make delivery economically worthwhile for pharmacies and affordable for clients:

> [The goal should be] to subsidize [the cost of] their [injectable PrEP] products to almost zero. That way, pharmacies do not have to pass all of the cost [on] to a client. They can … charge [clients] for the service [i.e., the pharmacy provider’s time], but not for the products. *(Female stakeholder from pharmacy professional society, in their 30s)*

**Table 4.** Pharmacy supplies and pharmacy provider competencies needed for injectable PrEP delivery.

| Pharmacy Supplies | Pharmacy Provider Competencies |
| --- | --- |
| • HIV testing kits<br>• Injectable PrEP drug<br>• Syringes<br>• Alcohol swabs/sanitizer<br>• Gloves<br>• Sharps disposal box and bins for biohazardous waste<br>• Exam table or couch<br>• Logbook<br>• Airtime for following up with clients<br>• Informational brochures for clients<br>• Job aids (e.g., provider handbook) | • Injectable PrEP guidelines<br>• Proper drug storage<br>• Counseling on HIV prevention options, including multiple PrEP products<br>• HIV testing and counseling<br>• Injection administration (theory and practice)<br>• Biohazardous waste disposal<br>• Handling adverse reactions<br>• Side effects monitoring<br>• Pharmacovigilance<br>• PrEP specialist consultation and referrals |

### Individuals domain

#### Client need

Participants felt that availing injectable PrEP at pharmacies would fulfill a critical need among clients for HIV services that are accessible, fast, private, and respectful. Accessibility was largely viewed as a byproduct of pharmacies’ proximity to where people live and work and their long operating hours. Fast service times were attributed to pharmacies’ short queues and few, if any, hand-offs between providers. Privacy would be protected, in part, by the fact that pharmacies offer a broad array of products and services for a wide range of health needs (not specifically HIV) and by the injectable PrEP delivery occurring in a nondescript room, precluding other customers from knowing what service the client received. Lastly, a few participants asserted that pharmacy providers generally treat clients with more patience and kindness than their overworked, clinic-based counterparts and might, therefore, satisfy some clients’ need for respectful and nonjudgmental treatment.

#### Client capability

Low client knowledge about injectable PrEP and its availability at pharmacies were identified as potential barriers to service uptake, with participants recommending a variety of approaches to increase client awareness, including social media, TV, and radio ads; print materials, like posters and brochures; and in-person talks at clinics and community events.

#### Client opportunity

Participants said that clients’ opportunity to engage in pharmacy-delivered injectable PrEP services would be influenced by the cost of services as well as the quantity and distribution of pharmacies offering this service. Concerned about what pharmacies would charge for injectable PrEP, clients and stakeholders underscored the need for it to be “affordable.” Most clients thought injectable PrEP should be free or cost 100 Kenyan shillings (∼$1 USD) at most, anticipating that any higher cost would price out a large portion of the target population. A few stakeholders felt that, if the government supplied pharmacies with the injectable PrEP drug for free, it could impose caps on how much, if anything, pharmacies charge clients. Several clients and providers also noted that, if injectable PrEP were only available at a handful of pharmacies, clients who travel frequently for school or work would have difficulty returning for follow-up doses; they, therefore, proposed making injectable PrEP available at pharmacies across the country.

#### Client motivation

Participants predicted that clients’ motivation to engage in pharmacy-delivered injectable PrEP services would be influenced by their trust in the drug and confidence in the provider’s integrity and competency. For the former, participants felt that, given injectable PrEP’s newness, it would be especially important to assure people that the drug is safe, effective, and government-approved. For the latter, participants warned of “quacks”—unlicensed or underqualified individuals who provide poor quality services—and asserted that clients would only be willing to get injectable PrEP from a provider whom they trusted would maintain confidentiality and provide genuine (not counterfeit) drugs:

> [At some pharmacies] they may lie that they have given you that medication, but maybe it’s not the [right] one. They should show us the medicine before so that I can see the [expiration] date … [and ensure] they don’t inject me if it’s expired. *(Female pharmacy PrEP client who did not refill, in their 30s)*

Participants additionally thought that clients would consider whether the provider understands the injection and is capable of safe injection administration:

> You have to find out if the person giving you the injection is well knowledgeable in that field, or are they just administering it for the sake [of making money]? … Someone who is qualified will not harm you. *(Female pharmacy PrEP client who refilled, in their 20s)*

To help clients identify qualified injectable PrEP providers, some participants recommended that providers receive and display a certificate of training. Concerning the professional cadre, about half of clients thought they would be comfortable getting CAB-LA from a trained pharmacy provider, whereas the remainder anticipated they would prefer a nurse or doctor. About one-third of clients specified they would only get lenacapavir from a doctor or nurse in a clinic setting because they perceived the risk of harm to be greater given the abdominal injection site. Lastly, participants said some clients—especially those who fear judgement of their sexual activity— would take into account the provider’s bedside manner:

> I would consider the pharmacy provider I am going to [and] if it is someone who understands me. … It is important to speak to clients well and encourage them … [and not] start telling them, “What you are doing is not good. Even if you are using PrEP, you should not [have multiple sex partners].” *(Female clinic PrEP client, in their 20s)*

### Provider capability

Most participants felt as though, with proper training and support, pharmacy providers would be capable of safely delivering this service. **Table 4** summarizes the knowledge, skills, and competencies that participants thought pharmacy providers would need to deliver injectable PrEP. Providers anticipated that administering lenacapavir would be easy, noting that it is not uncommon for pharmacy providers to give subcutaneous injections after receiving on-the-job training and that some have already been trained by the MOH to deliver injectable contraception. A handful of participants said the government could consider requiring pharmacies to have nurses and/or doctors on staff to administer the injection, but that acquiring such staff would be cost-prohibitive for most pharmacies.

### Provider opportunity

Nearly all providers expressed concern that the amount of time it takes to deliver injectable PrEP would exceed what pharmacy providers typically have available. Most of those with prior experience delivering oral PrEP said it was time-consuming and especially challenging during peak business hours. These providers worried that counseling clients on multiple forms of PrEP—oral, CAB-LA, and/or lenacapavir—would be even more time-consuming, and a few anticipated they would push clients to pick the form that was fastest for them to deliver. Some felt lenacapavir—with only two visits per year—would be more feasible for pharmacy providers to deliver. Participants suggested several ways to mitigate barriers to provider availability, including pharmacies having two providers per shift; delivering injectable PrEP by appointment; and the MOH seconding staff to pharmacies.

### Provider motivation

Participants anticipated that the primary motivator for pharmacy providers to deliver injectable PrEP would be the prospect of financial gain. These participants pointed out that having a provider attend to injectable PrEP clients in a back room could lead to missed sales—and lost profit—for the pharmacy. A few providers further noted that, in some pharmacies, the owner incentivizes employees by paying bonuses to those who hit daily sales targets. These participants thought this compensation structure might disincentivize providers from delivering injectable PrEP, as many other products take less time to deliver and fetch higher profits:

> “Here [at this pharmacy], we work with [sales] targets. Everyone has their targets, so even if I am serving the client here [in the back room], I must make sales. If they [injectable PrEP clients] consume your time here, the more you are losing there. … The big challenge [with delivering oral PrEP] was the fact that it was time-consuming.” *(Female pharm tech with prior PrEP delivery experience, in their 30s)*

To mitigate this challenge, a few participants recommended the government or implementer pay a direct incentive to the provider delivering the service, rather than compensating pharmacy owners and leaving it up to them to decide how much, if anything, to share with the provider.

## DISCUSSION

In this qualitative study, Kenyan oral PrEP users, pharmacy providers, and other key stakeholders were generally supportive of the idea of pharmacy-delivered injectable PrEP and expressed cautious optimism that, with the right support and oversight, this delivery channel could increase injectable PrEP access. Participants identified several characteristics of pharmacies that might facilitate injectable PrEP implementation, such as long operating hours; proximity to where people live and work; discrete service delivery; and lack of HIV stigma. Participants also identified barriers at multiple levels that could hinder the acceptability, feasibility, and/or adoption of injectable PrEP among pharmacy clients and providers. Findings from this research can inform decision-making about whether and how to expand injectable PrEP services to private pharmacies, including selection of implementation strategies that might best position this delivery channel for success in Kenya and similar settings.

Although many of the anticipated barriers identified in this formative research echo those reported in other injectable PrEP studies and real-world programs, ^17, 37–41^ a subset of these barriers are likely to be more pronounced in private pharmacies than in traditional healthcare delivery channels.^21^ These potential “deal breakers” for pharmacy delivery include gaps in enabling pharmacy policy; exclusion of private pharmacies from injectable PrEP supply chains and health information systems; disincentives for private pharmacy providers to dedicate time to injectable PrEP delivery; and clients’ general wariness of the private pharmacy sector, which has historically been plagued by unlicensed outlets with unqualified staff and/or counterfeit drugs.^42, 43^ In private pharmacies, providers face low provider-to-client ratios and pressure to make sales—a core mandate of any for-profit business. These circumstances could undermine the availability and quality of pharmacy-delivered injectable PrEP services, with busy providers turning clients away or cutting corners on time-consuming delivery components, like choice-based counseling.^44^ To mitigate this risk, implementation strategies that reduce the amount of time pharmacy providers spend on delivery, such as availing staff who can assist with delivery in person or via telehealth,^27,45^ should be considered. To build client trust in this delivery channel, other strategies will be necessary, such as amending public registries of licensed pharmacies and pharmacy providers^24^ to indicate whether the premise or person is authorized to deliver injectable PrEP, and directing clients to apps they can use at the point of delivery to verify the authenticity of injectable PrEP drugs.^46^

As a later-adopter of injectable PrEP, Kenya can capitalize on information that has already been generated about delivery in other settings. For example, several other injectable PrEP research studies and real-world programs^17, 37–41^ have reported implementation barriers similar or identical to those anticipated by our study participants, such as low client knowledge of injectable PrEP and concerns about its safety; client travel as a barrier to attending follow-up visits; and gaps in provider knowledge. Kenya could potentially leapfrog these common barriers by taking into consideration the implementation strategies that have worked well in other settings and leveraging and adapting existing resources for the pharmacy context, such as information, communication, and education materials;^17^ provider training curricula;^47–50^ job aids;^51^ and documentation and reporting tools.^21, 52^ Importantly, several major implementation barriers that did not come up in this formative research have emerged in other settings and warrant careful consideration, such as type of HIV testing to be used at CAB-LA initiation and during CAB-LA use to identify acute infections or long-acting early viral inhibition (LEVI) syndrome,^53^ and how to ensure that clients who return late for CAB-LA re-doses (or who discontinue CAB-LA) use oral PrEP during delays (or after discontinuing) to reduce the risk of acquiring drug-resistant HIV.^54^

In **Table 5**, we outline two example pathways for implementing injectable PrEP services at private pharmacies in Kenya in the short and long term. For expedited implementation in the short term, we suggest developing injectable PrEP certification programs for pharmacy providers; granting special permission to pharmacies that already have PrEP prescribers on staff; and seconding PrEP prescribers to pharmacies. For sustained implementation in the long term, we suggest revising national policies and guidelines (e.g., PrEP implementation guidelines, pharmacy providers’ scope of practice); integrating pharmacies into Kenya’s health information system; and amending public registries so clients can identify pharmacies and providers authorized to deliver injectable PrEP. Of note, these pathways are not mutually exclusive and could be pursued concurrently, especially since policy changes could take years to achieve.

**Table 5.** Potential actions to address barriers specific to pharmacy-delivered injectable PrEP for sustained and expedited implementation.

| Barrier to pharmacy delivery<sup>1</sup> | Example actions |  |
| --- | --- | --- |
|  | Long-term pathway for sustained implementation at scale | Short-term pathway for expedited implementation |
| **Policy** |  |  |
| **Gaps in enabling policies** hinder legal delivery of injectable PrEP at pharmacies | <u>Kenya Ministry of Health (MOH):</u><br>Develop **national injectable PrEP guidelines** specifying which cadres of healthcare professional can deliver injectable PrEP.<br>Revise **essential medicines list** to indicate that injectable PrEP can be delivered at pharmacies meeting specific criteria.<br>Revise **scope of practice of pharmacists and/or pharm techs** to include requisite injectable PrEP competencies (e.g., permission to administer subcutaneous and/or intramuscular injections; prescribing privileges with remote clinician supervision). | <u>Kenya MOH:</u><br>Grant **special permission** to pharmacies with PrEP prescribers<sup>2</sup> on staff to initiate and manage clients on injectable PrEP.<br>**Second PrEP prescribers<sup>2</sup>** to select pharmacies (e.g., in HIV hot spots). Create an **injectable PrEP certificate program** for pharmacy providers that confers permission to initiate and manage clients on injectable PrEP under <u>Kenya MOH, implementing partners, and civil society organizations</u>:<br>remote clinician supervision. |
| **Procurement and financing** |  |  |
| Private pharmacies are **not currently part of the Kenya's PrEP supply chain system** or pooled procurement mechanisms | <u>Kenya MOH:</u><br>Integrate pharmacies into the **national PrEP supply chain** (i.e., issue them codes for ordering injectable PrEP directly from government warehouses).<br>**Determine compensation** to private pharmacies (and/or allow pharmacies to charge clients) for injectable PrEP services, ensuring that the prospective profit is commensurate with delivery costs incurred.<br>*[If applicable]* **Establish a mechanism to pay pharmacies** for injectable PrEP services rendered. | <u>Kenya MOH:</u><br>Link **pharmacies to nearby public health facilities** to obtain PrEP drugs and ancillary supplies (e.g., HIV testing kits, needles, sharps disposal containers) from the national stock.<br>**Compensate pharmacies** and limit what they can charge clients for injectable PrEP services. |
| **Health information (HIS) systems** |  |  |
| **Lack of interoperability** between Kenya's national HIS and those used by private pharmacies. | <u>Kenya MOH:</u><br>**Establish minimum criteria for HIS systems** for injectable PrEP delivery.<br>Identify specific HIS systems that could be used for injectable PrEP delivery, including for delivery at pharmacies. | <u>Kenya MOH</u><br>**Implement the Kenya EMR PrEP module**, currently used at select public health facilities offering CAB-LA, at private pharmacies. |
| **Pharmacy provider availability and motivation** |  |  |
| Private pharmacies' **profit-generation focus** and **low staffing levels** | <u>Kenya MOH and/or implementing partners:</u><br>Explore **alternative financial arrangements** (e.g., pay-for-performance) that might motivate delivery of high-quality injectable PrEP services.<br>**Create legal pathways**, as needed, for strategies found to reduce burden on pharmacy provider time (e.g., telehealth scripting; HIV self-testing).<br>**Adapt/simplify injectable PrEP reporting tools** for the pharmacy setting. | <u>Kenya MOH and/or implementing partners:</u><br>**Second staff** to pharmacies to assist with delivery in person or remotely.<br>Consider **direct financial incentives** to individual pharmacy providers delivering services.<br>Test **implementation strategies** that could reduce burden on pharmacy provider time (e.g., option to self-screen for HIV risk; HIV self-testing with optional provider assistance; client-facing decision support tools). |
| **Public confidence in private pharmacy sector** |  |  |
| Client concerns about **pharmacy providers' competency** and/or drug authenticity; **public mistrust** of the private pharmacy sector | <u>Kenya MOH and implementing partners:</u><br>**Amend existing public registries** of pharmacies and pharmacy providers to indicate which are authorized to deliver injectable PrEP.<br>**Create formal partnerships** between private pharmacies and public health facilities. | Create a **temporary registry** of pharmacy providers who have completed injectable PrEP training and issue **them training certificates** (to display in pharmacy) that clients can verify (e.g. via QR code).<br>Include in demand generation materials **information about which pharmacies and providers are authorized** to offer injectable PrEP and steps clients can take to verify this information at the point of delivery. |

<sup>1</sup>These barriers are informed by, but not limited to, those reported by study participants and detailed in Tables 2 and 3.
<sup>2</sup>Clinical officers or nurses working under remote clinician supervision

Our study has limitations. First, our purposively selected sample is not representative of all PrEP stakeholders in Kenya (e.g., adolescents; donor agencies); as such, we likely did not capture all relevant implementation determinants, and our findings may have limited generalizability to other individuals and settings. Second, we provided participants with standardized descriptions of CAB-LA and lenacapavir to ensure their responses were anchored to a shared understanding of basic product facts; however, unmeasured factors—such as baseline knowledge about different types of injections—may have influenced participant responses, especially client concerns about lenacapavir’s abdominal injection site. Future research should consider using visuals or demos on anatomical models. Lastly, because pharmacy-delivered injectable PrEP is hypothetical, participant perspectives may not accurately reflect how they would feel about the innovation in a real world scenario.

## CONCLUSION

Key stakeholders of HIV prevention in Kenya are cautiously optimistic that private pharmacies could be an effective delivery channel for injectable PrEP. Successful implementation will require integrating private pharmacies into multiple subsystems of Kenya’s public health sector and proactively addressing multi-level barriers. If private pharmacies facilitate access to and use of injectable PrEP, they could play a pivotal role in ending HIV as a public health threat.

## STATEMENTS AND DECLARATIONS

### ETHICS APPROVAL

This study was approved by the Institutional Review Board of the University of Washington (STUDY00016809) and the Kenya Scientific Ethics Review Unit subcommittees for the Center for Microbiology Research (protocol number: CMR/P00239-01-2023/4706) and Center for Clinical Research (protocol number: CCR/0308/4709).

### CONSENT TO PARTICIPATE

Informed consent was obtained from all individual participants included in this study.

## ACKNOWLEDGEMENTS

The team gratefully acknowledges the individuals who contributed to this research as study participants and the Kenya National AIDS and STI Control Programme for their collaboration.

## DATA AVAILABILITY STATEMENT

Data are available upon reasonable request.

