## Appendix A for "Implementing long-acting injectable HIV pre-exposure prophylaxis services at private pharmacies in Kenya: Client, pharmacy provider, and key stakeholder perspectives on potential challenges and opportunities"

**Electronic supplementary materials for**

**Appendix A. Interview guides**

**In-Depth Interview Guide: Pharmacy Clients**

**Interviewer instructions: Administer informed consent. Once signed, begin guide.**

**0.0 Interview Information**

***Fill out items A through I prior to starting interview.***

(a) **Informed consent has been administered**: YES	/	NO

*If consent form has not been signed by participant,*

*interview must not proceed.*

(b) **Client PTID from Pilot Extension Study**: _____________________________
(c) **Interviewee category** *(according to study records; select one):*

*☐* Pharm PrEP Naive: Has used PrEP before but never from a study pharmacy

*☐* PrEP Refiller: Refilled PrEP at study pharmacy at least once

*☐* PrEP Discontinuer: Initiated PrEP at study pharmacy but never refilled there and indicated in post-study survey that they did not wish to get PrEP at clinic.

(d) **Interview ID**: _____________________________

*Format: Interviewee Type-DDMMYY-Number interview conducted that day*

*where “CR” = Client Refiller; “CD” = Client Discontinuer; and “CN” is “Client Naive”*

*(e.g., “CR-150423-02” represents the second Client Refiller interviewed on 15 Apr 2023)*

(e) **Date of interview**: ____/_____/_______

*Format: DD/MM/YYYY*

(f) **Location of interview**: __________________________________________

(g) **Interviewer’s full name**: _________________________________________

(h) **Interview start time**: ____________________

*Format: HH:MM am or pm*

(i) **Interview end time**: _____________________

*Format: HH:MM am or pm*

**Facilitator introduction: [DO NOT READ; GUIDE ONLY]**

Hello. My name is _______, and I am a ________, working at _______. Thank you for taking the time to talk with me today.

The purpose of this interview is to understand your past experiences, if any, of getting PrEP services at a pharmacy and your thoughts about new forms of PrEP that may be offered at pharmacies in the future.

There are no right or wrong answers to these questions. People have different views and we are interested to learn more about these experiences from you. Today, you are in the role of a teacher and I am here to learn from you since you are an expert in your own life experiences and opinions.

This interview should take around an hour to complete. Please let me know if at any time you have questions, if something I say is not clear, or if you need to take a break.

| 1. **[*INTERVIEWER: Mark the participant’s sex.*]** | *☐ Male*  *☐ Female* |
| --- | --- |
| ***START INTERVIEW HERE.*** | |
| I’d like to start off by asking you some basic questions about yourself. | |
| 2. How old are you? | _________ years old |
| 3. What is the highest **level** of school you have attended? *(Select one)* | *☐ No schooling*  *☐ Pre-primary*  *☐ Primary*  *☐ Secondary (vocational, ‘A’ level’, etc.)*  *☐ University/College (including post-secondary vocational school)*  *☐ Prefer not to answer* |
| 4. In total, how many years did you attend school? | ______ years total  *☐ Prefer not to answer* |
| 5. Which of the following best describes your current occupation? *(Select one)* | *☐ No income*  *☐ Student*  *☐ Professional*  *☐ Trade/sales*  *☐ Laborer/semi-skilled*  *☐ Farming/animal raising*  *☐ Other: _____________________________* |
| 6. About how much money do you make per month? | _______ KSH per month |
| 7. What is your current marital status? *(Select one)* | *☐ Married*  *☐ Not married*  *☐ Prefer not to answer* |
| 8. What is your current relationship status? *(Select one)* | *☐ Single, no partners*  *☐ Casual partner(s) only*  *☐ One primary partner only*  *☐ One primary partner and casual partners*  *☐ Other: _____________________________*  *☐ Prefer not to answer* |
| 9. Do you have any children? | *☐ Yes*  *☐ No* |
| 10. **[If Q9=YES]** How many children do you have? | *__________* children |
| 11. Have you ever gotten PrEP at a clinic? | *☐ Yes*  *☐ No* |
| 12. **[If Item C= “PrEP Refiller” or “PrEP discontinuer” & If Q11=YES]** Did you get PrEP at a clinic *before* getting it at the pharmacy or *after*? | *☐ Before (i.e., before the Pilot Extension study started)*  *☐ After (i.e., after the Pilot Extension study ended)*  *☐ Both before and after* |

**TOPIC 1: Experience getting oral PrEP at the pharmacy**

**[For PrEP Refillers & PrEP Discontinuers only]**

Part of the reason why we were interested in interviewing you was because you previously got PrEP from a pharmacy.

As you may recall, in order to get PrEP at that pharmacy, the pharmacy provider asked you some questions about your sexual behaviors and health and helped you do an HIV test to confirm that you were HIV-negative. Then the provider gave you one bottle of PrEP pills.

I’m interested to hear about your experience getting PrEP at the pharmacy. Let’s start with the positives.

Was there anything that you **liked** about getting PrEP at the pharmacy?

*[If not covered in their response to the above:]*

- How easy or hard was it for you to get yourself to the pharmacy for PrEP?

*[Probe, if needed]*

- How close is the pharmacy from where you live or work?
- Was the pharmacy usually open when you wanted to go there?
- While getting PrEP at the pharmacy, how comfortable or uncomfortable did you feel?

*[If not covered in response to the above:]*

- While answering questions about your sexual behaviors and doing the HIV test, did you feel like there was enough privacy? Why or why not?
- What sex was the pharmacy provider? About how old do you think [he/she] was? How comfortable were you discussing your sexual behaviors with a [male/female] provider in their [20s/30s/40s/50s/60s]? Would you have preferred to talk to someone different? Please explain.
- How confident were you that the pharmacy provider would keep your information confidential and not tell it to anyone else?
- In general, do you feel like the PrEP services you received at the pharmacy were good quality? Why or why not?

- Tell me about anything in particular that made you want to get PrEP at a pharmacy rather than somewhere else, like a clinic?
  - *[If client has ever gotten PrEP at a clinic]* How does your experience getting PrEP at the pharmacy compare to getting it at a clinic?

*[If in response to previous questions, the interviewee mentioned 1+ thing that they disliked about getting PrEP at the pharmacy]* Ok. So it sounds like there were a few things that you did **not** like about getting PrEP at the pharmacy, such as *[dislikes previously mentioned, e.g., “the fact that the pharmacy only had a male provider”]*. Is there anything else that you did **not** like about getting PrEP at the pharmacy?

*[If interviewee hasn’t mentioned any dislikes prior to this point]* Was there anything that you **disliked** about getting PrEP at the pharmacy?

*[Probe, as needed, to understand if the dislike is specifically related to the pharmacy or to PrEP itself, like not liking the side effects, not liking only getting 1 bottle of PrEP at initiation, etc.]*

*[If not covered in their response to the above:]*

- What would you change about how PrEP is delivered at the pharmacy? Please explain.

*[Probe, as needed, to understand what exactly the interviewee is proposing and what they think would improve if this change were implemented (e.g., would it make pharmacy PrEP services more convenient/accessible, more private, less stigmatizing, etc.)?*

**[TO ASK PrEP DISCONTINUERS ONLY]**

After initiating PrEP at the pharmacy, you did not return to refill your PrEP prescription, and you reported in our post-study survey that you had stopped taking PrEP primarily because you didn’t want to get it at a clinic.

*[If not covered already]* Can you tell me more about why you **didn’t** return to the pharmacy to refill your PrEP prescription?

*[Probe, as needed, to understand if their reasons for discontinuing were specific to the pharmacy setting, e.g., not feeling comfortable HIV testing at the pharmacy; the pharmacy being inconvenient for them to go to; etc.]*

And can you tell me more about why, after the study ended, you **didn’t** want to continue getting PrEP at a clinic?

*[Probe, if needed:]*

- Do you know of any clinics that offer PrEP services?
  - *[If yes]* Are these clinics public, private, or a mix?
- How easy or hard do you think it would be for you to get yourself to a clinic for PrEP?

*[Probe, if needed]*

- Is there a clinic near where you live or work that has PrEP?
- *[If yes]* Is this clinic generally open when you need it to be?
- How comfortable or uncomfortable do you think you would be getting PrEP at a clinic? Please explain.
- In general, do you think a clinic would offer good quality PrEP services? Why or why not?

**[TO ASK PHARM PrEP NAÏVE CLIENTS ONLY]**

Part of the reason why we were interested in interviewing you is because you previously got PrEP from a clinic. I’ll start off by asking you some basic questions about your experience using PrEP.

When did you first start taking PrEP?

*[Probe, as needed, to understand if client has taken PrEP continuously, taken breaks and then restarted, etc.]*

Are you still taking PrEP today?

In total, about how many months have you taken PrEP?

Where do you—or did you use to—get PrEP from?

*[Probe, as needed, to understand if client gets/got PrEP from a public or a private facility, from multiple types of facilities, etc.]*

*[If client indicates that they are no longer taking PrEP]*: Why did you stop taking PrEP?

I’d like you to imagine that you had the option to get PrEP at a private pharmacy, rather than having to go to a clinic. To get PrEP at a pharmacy, the chemist would ask you some questions about your health and your sexual behaviors to see if you are at risk of getting HIV. The chemist would also test you for HIV, and if they found you eligible for PrEP, they would dispense you PrEP pills. You would return to the pharmacy for follow-up visits and refills.

How do you feel about the idea of PrEP being available at the chemist?

*[Probe, if needed*: What are some of the potential advantages of getting PrEP at a pharmacy? What are some of the potential disadvantages?*]*

**TOPIC 2: CAB PrEP**

I’m now going to talk to you about a new form of PrEP called “cabotegravir PrEP” or “CAB PrEP”. It is also sometimes called “injectable PrEP”.

Have you ever heard of “CAB PrEP” or “injectable PrEP” before?

*[If yes]*

- Where did you hear about it?
- What have you heard about it?

Ok. I’m going to tell you a little bit about CAB PrEP *[Hand interviewee a printed copy of this description so they can follow along]:*

**Description of cabotegravir PrEP (CAB PrEP) or “injectable PrEP”**

- CAB PrEP is a new form of PrEP that is not yet available in Kenya. It is currently being reviewed by the Kenya Ministry of Health.
- Like the PrEP pills you have taken, CAB PrEP is used to **prevent HIV**. Like PrEP pills, CAB PrEP is used by people who think they might get exposed to HIV in the near future, and in order to work, CAB PrEP must be taken **before** being exposed to HIV.
- Instead of taking one pill every day, people using CAB PrEP get **one injection** **every two months**. This injection contains a drug called cabotegravir that helps prevent HIV. The injection goes into the **upper part of the bum/buttocks**.
- Like the PrEP pills you have taken, CAB PrEP is more than 90% effective at preventing HIV in the whole body, regardless of how one is exposed to HIV.
- Like PrEP pills, CAB PrEP does **not** protect against pregnancy or sexually transmitted infections. So if a person using CAB PrEP wants to protect against pregnancy and/or infections like chlamydia and gonorrhea, they would still need to use condoms and/or contraception.
- Before each CAB PrEP injection, one must get tested for HIV to confirm that they are HIV-negative.
- Side effects from CAB PrEP are usually limited to the injection site (the place on the bum where the injection goes in). This area may be a little tender, bruised, or swollen for a couple of days after getting the injection.

If you felt like you were at risk of getting HIV, do you think you would be interested in getting a CAB PrEP injection every two months? Why or why not?

*[If yes, probe, as needed, to understand what they think the benefits of CAB PrEP would be over PrEP pills. If no or unsure, probe, as needed, to understand their concerns. For example, “In general, how do you feel about getting an injection versus taking a pill?”]*

Is there any additional information about CAB PrEP that you would want to know before deciding whether to take it? Please explain.

I’d like you to imagine that the Ministry of Health has approved CAB PrEP and that you **are** interested in starting (or switching to) CAB PrEP.

Would you want CAB PrEP **to be available** at pharmacies? Why or why not?

*[If interviewee says “no”, probe, as needed, to understand the interviewee’s rationale. For example, ask, “What do you think would happen if CAB PrEP were made available at pharmacies?” to see if they have concerns about it being prohibitively expensive, about pharmacy providers’ ability to safely administer injections, about CAB PrEP being misused/abused, etc.]*

If CAB PrEP were available at pharmacies, do you think you would be **willing to get it there**?

- *[If “no” or “it depends”]*
  - Why not?
  - Under what circumstances, if any, do you think you would be willing to get CAB PrEP at a pharmacy? *[Tailor probes based on prior responses. For example, if interviewee expressed concerns about a pharmacy provider giving the injection, then ask: “If the pharmacy had a nurse or doctor administering the injection, would that change your mind?” If interviewee expressed concern about showing their bum to a pharmacy provider of a particular sex, ask: “If the pharmacy provider giving you the injection were a [male/female], would that change you mind?” If interviewee expressed concern about cost, ask: “Would you be willing to get CAB PrEP at a pharmacy if it were free?”]*
- *[If yes]*
  - How comfortable would you feel about getting the injection from a trained pharmacy provider?
    - Who would you want to administer the injection?
  - Would you feel comfortable getting the injection in a private back room of the pharmacy?
  - Are there any other conditions a pharmacy would have to meet in order for you to be willing to get CAB PrEP there?

If CAB PrEP were available at pharmacies, do you think **other people with HIV risk** would be **willing to get it there**? Why or why not?

*[Probe, as needed, to understand if they are thinking of a specific population—e.g., young people, married people, sex workers, MSM—and, if so, why they think this population would or wouldn’t be willing to get CAB PrEP at a pharmacy.]*

What, if anything, might a chemist do to make clients **feel more comfortable** getting CAB PrEP at the pharmacy? *[Probe, as needed, to understand if the interviewee is recommending changes to the physical space/furniture/equipment, how the pharmacy provider interacts with clients, how the pharmacy or ministry of health promotes CAB PrEP in the community, etc.]*

If CAB PrEP were available at pharmacies, what do you think would be the best way to **spread the word** about its availability there? *[Probe, as needed, to understand recommended platform, target audience, and/or key points to include in messaging.]*

**TOPIC 3: 6-month PrEP Injection**

I’d like to tell you about another type of PrEP injection that is currently in being investigated in clinical trials. *[Hand interviewee a printed copy of this description so they can follow along]:*

**Description of 6-month PrEP injection:**

- There is another form of injectable PrEP that is **not yet available to the public** anywhere in the world.
- Early findings from research studies suggest that this injection might prevent HIV with just **two doses per year** (once every 6 months).
- This injection contains a drug called lenacapavir that helps prevent HIV. The injection goes into the **abdomen**.
- Before each injection, one must get tested for HIV to confirm that they are HIV-negative.

I’d like you to imagine that the clinical trials prove that the 6-month PrEP injection is as effective at preventing HIV as PrEP pills and CAB PrEP.

If you felt like you were at risk of getting HIV, do you think you would be interested in getting a 6-month PrEP injection twice per year to prevent HIV? Why or why not?

*[If yes, probe, as needed, to understand what they think the benefits of a 6-month PrEP injection would be over CAB PrEP and PrEP pills. If no or unsure, probe, as needed, to understand their concerns.”]*

Whereas CAB PrEP injections are in the upper part of the bum/buttocks, this 6-month PrEP injection goes into the abdomen. Do you think this would affect your willingness to get the 6-month PrEP injection? Please explain.

Is there any additional information about a 6-month PrEP injection that you would want to know before deciding whether to take it?

You’ve already expressed to me your feelings about CAB PrEP being delivered in pharmacies. Do you feel any differently about the idea of a 6-month PrEP injection being delivered in pharmacies? Please explain.

*[If yes, probe, as needed, to understand what they feel differently about and why. For example, if interviewee previously said they were in favor of CAB PrEP being available in pharmacies but is now saying they don’t think a 6-month PrEP injection should be in pharmacies, ask why)].*

**TOPIC 4 [only for female interviewees]: Dapivirine vaginal ring**

I’m now going to talk to you about another new form of PrEP called the dapivirine vaginal ring or “PrEP ring”.

Have you ever heard of the dapivirine ring or “PrEP ring” before?

*[If yes]*

- Where did you hear about it?
- What have you heard about it?

Ok. I’m going to tell you a little bit about the PrEP ring *[Hand interviewee a printed copy of this description so they can follow along]:*

**Description of dapivirine vaginal ring or “PrEP ring”**

- The PrEP ring is a new form of PrEP that was recently approved by the Kenya Ministry of Health and is being introduced at public health facilities.
- Like the PrEP pills you have taken, the PrEP ring is used to **prevent HIV**. The PrEP ring is used by women who think they might get exposed to HIV during **vaginal sex** in the near future, and in order to work, the PrEP ring must be used **before** being exposed to HIV.
- The PrEP ring is made of flexible silicone rubber and contains a drug called *dapivirine* that helps prevent HIV.
- A woman can insert the PrEP ring into her vagina by herself and leave it in for 28 days. During these 28 days, the ring does not have to be taken out or cleaned, even after sex or menstruation. At the end of 28 days, she can take the ring out by herself and insert a new ring.
- The PrEP ring is about 50% effective at preventing HIV in the vagina. It does not prevent HIV in other parts of the body, such as the anus.
- Like PrEP pills, the PrEP ring does **not** protect against pregnancy or sexually transmitted infections. So if a woman using the PrEP ring wants to protect herself against pregnancy and/or infections like chlamydia and gonorrhea, she would still need to use condoms and/or contraception.
- As with PrEP pills, women using the PrEP ring would need to periodically get tested for HIV to ensure she is still HIV-negative.
- Side effects from the PrEP ring are uncommon. Some users report very mild side effects, like vaginal discomfort, that usually go away after a few days.

I’d like you to imagine that that you **are** interested in using the PrEP ring. Please also imagine that, in order to get the PrEP ring, you would have to answer questions about your sexual behaviors (to confirm that you have HIV risk), undergo an HIV test (to confirm that you are HIV-negative), and get counseled on how to insert the PrEP ring into the vagina. You would then be given a PrEP ring to take home with you to insert yourself or, if you wish, you could have a healthcare provider help you insert the PrEP ring.

Would you want the PrEP ring **to be available** at pharmacies? Why or why not?

*[If interviewee says “no”, probe, as needed, to understand the interviewee’s rationale. For example, ask, “What do you think would happen if the PrEP ring were made available at pharmacies?” to see if they have concerns about it being prohibitively expensive, about privacy at pharmacies, etc.]*

If the PrEP ring were available at pharmacies, do you think you would be **willing to get it there**?

- *[If “no” or “it depends”]*
  - Why not?
  - Under what circumstances, if any, do you think you would be willing to get the PrEP ring at a pharmacy? *[Tailor probes based on prior responses. For example, if interviewee expressed concerns about a pharmacy provider instructing them on how to use the PrEP ring, then ask: “If the pharmacy had a nurse or doctor to counsel about the PrEP ring and assist with insertion, as needed, would that change your mind?” If interviewee expressed concern about the pharmacy provider being a particular sex, ask: “If the pharmacy provider giving you the PrEP ring were a [male/female], would that change you mind?” If interviewee expressed concern about cost, ask: “Would you be willing to get the PrEP ring at a pharmacy if it were free?”]*
- *[If yes]*
  - How comfortable would you feel about getting the PrEP ring from a trained pharmacy provider?
  - Who would you want to give you the PrEP ring?
  - Would you feel comfortable getting counseled about the PrEP ring in a private back room of the pharmacy?
  - After using a ring, would you feel comfortable throwing it away in the trash?
  - Are there any other conditions a pharmacy would have to meet in order for you to be willing to get the PrEP ring there?

If the PrEP ring were available at pharmacies, do you think **other women with HIV risk** would be **willing to get it there**? Why or why not?

*[Probe, as needed, to understand if they are thinking of a specific population—e.g., young women, unmarried women, female sex workers,—and, if so, why they think this population would or wouldn’t be willing to get the PrEP ring at a pharmacy.]*

What, if anything, might a chemist do to make women **feel more comfortable** getting the PrEP ring at the pharmacy? *[Probe, as needed, to understand if the interviewee is recommending changes to the physical space/furniture/equipment, how the pharmacy provider interacts with clients, how the pharmacy or ministry of health promotes the PrEP ring in the community, etc.]*

If the PrEP ring were available at pharmacies, what do you think would be the best way to **spread the word** about its availability there? *[Probe, as needed, to understand recommended platform, target female audience, and/or key points to include in messaging.]*

**TOPIC 5: Closing thoughts**

Is there anything else you’d like to share about your experience getting PrEP pills at the pharmacy or ways you think pharmacies might succeed at delivering CAB PrEP [and the PrEP ring]?

We’ve covered all the topics I wanted to discuss today. Thanks very much for your time. I’m going to shut off the recorder now.

If you have any additional questions about CAB PrEP [or the PrEP ring], I’d be happy to answer those or tell you where you can find more information.

**[Mark interview end time on page 1 (Item I).]**

**In-Depth Interview Guide: Pharmacy Providers**

**Interviewer instructions: Administer informed consent. Once signed, begin guide.**

**0.0 Interview Information**

***Fill out items A through I prior to starting interview.***

(j) **Informed consent has been administered**: YES	/	NO

*If consent form has not been signed by participant,*

*interview must not proceed.*

(k) **Interview ID**: _____________________________

*Format: Interviewee Type-DDMMYY-Number interview conducted that day*

*where “PE” = Provider Experienced and “PN” = Provider Naive*

*(e.g., “PE-150423-02” represents the second Pilot Extension pharmacy provider interviewed on 15 Apr 2023)*

(l) **Date of interview**: ____/_____/_______

*Format: DD/MM/YYYY*

(m) **Location of interview**: __________________________________________

(n) **Interviewer’s full name**: _________________________________________

(o) **Interview start time**: ____________________

*Format: HH:MM am or pm*

(p) **Interview end time**: _____________________

*Format: HH:MM am or pm*

**Facilitator introduction: [DO NOT READ; GUIDE ONLY]**

Hello. My name is _______, and I am a ________, working at _______. Thank you for taking the time to talk with me today.

The purpose of this interview is to understand your experiences or thoughts about delivering PrEP services at a pharmacy. We’d also like to understand your thoughts about potentially delivering new forms of PrEP that may become available in Kenya in the near future.

There are no right or wrong answers to these questions. People have different views and we are interested to learn more about these experiences from you. Today, you are in the role of a teacher and I am here to learn from you since you are an expert in your own life experiences and opinions.

This interview should take around an hour to complete. Please let me know if at any time you have questions, if something I say is not clear, or if you need to take a break.

| 13. **[*INTERVIEWER: Mark the participant’s sex.*]** | *☐ Male*  *☐ Female* | |
| --- | --- | --- |
| ***START INTERVIEW HERE.*** | | |
| I’d like to start off by asking you some basic questions about yourself. | | |
| 14. How old are you? | _________ years old | |
| 15. Are you a pharmacist or a pharmaceutical technologist? | *☐ Pharmacist*  *☐ Pharmaceutical technologist* | |
| 16. What is the highest level of training you received in order to become a [pharmacist/ pharmaceutical technologist?] | *☐ University degree*  *☐ College Diploma*  *☐ Other: _________________________* | |
| 17. How many years did your training last? | *_________ years* | |
| 18. Which pharmacy did you work at during the Pilot Extension study? | *☐ N/A*  Kisumu Pharmacies  *☐ Glory*  *☐ Kibos Road*  *☐ Kisa Urban*  *☐ Lakeview*  *☐ Maseno*  *☐ Main* | Kiambu Pharmacies  *☐ Cherathimo*  *☐ HealPlus*  *☐ Karangatha*  *☐ Makongeni*  *☐ Medihelp*  *☐ Sanaa* |
| 19. Do you own [*pharmacy selected for Q6 or, if Q6=NA, the pharmacy they currently work in*]? | *☐ Yes*  *☐ No* | |
| 20. How many years have you [worked at/owned] [*pharmacy selected for #6*]? | *_________ years* | |
| 21. How many years have you worked as a [pharmacist/ pharmaceutical technologist]? | *_________ years* | |
| 22. For how many months were you directly involved in PrEP delivery at [*pharmacy selected for #6*]?   *Include in this number any months spent delivering PrEP during the original pilot study, if applicable. Otherwise, if only participated in Pilot Extension, this number should not exceed 6.* | *☐ N/A*  ________ months | |

**TOPIC 1: Experience delivering oral PrEP at the pharmacy**

**[ONLY FOR PROVIDERS WHO PARTICIPATED IN THE PILOT EXTENSION STUDY]**

Part of the reason why we were interested in interviewing you is because you have experience delivering oral PrEP at a pharmacy.

As you know, when a client at your pharmacy was interested in PrEP, you had to ask them a series of questions to see if they were at risk of getting HIV and to ensure that they did not have any contraindications to PrEP, like a history of kidney disease. Then you also had to assist them with HIV self-testing and confirm that they were HIV-negative. And, lastly, you had to counsel them about PrEP and explain, for example, how it’s taken and why it is important to take PrEP every day for it to work.

Based on your experience, do you think pharmacies are a **good place to offer PrEP**? Why or why not?

*[If not covered in their response to the above:]*

- How **easy or hard** was it for you to deliver PrEP at the pharmacy?

*[If “hard”, probe, as necessary, to understand what, exactly, made it difficult. For example, if they talk about PrEP delivery taking too long, ask, “Which tasks related to PrEP delivery took the longest?” If they say they didn’t have enough knowledge/training, ask, “What kinds of things were you unsure about or didn’t feel like you knew how to do?”].*

- What **challenges** did you experience while delivering PrEP? Please explain.
  - *[If any challenges mentioned]* Did you find any ways to address these challenges? (Did you make any changes to how you do things at the pharmacy to mitigate these challenges?)
- Did you feel **comfortable talking about PrEP** to clients? *[If no]* What made you feel uncomfortable? *[Probe, as needed, to understand the drivers of this discomfort. For example, “Are you generally comfortable asking clients about their sexual behaviors?” “How did clients respond when you asked them if they were interested in learning more about a drug to prevent HIV?”, “Did you feel like you had sufficient knowledge to answer clients’ questions about PrEP?”]*

What, if anything, would make it **easier** for pharmacy providers like yourself to deliver PrEP at pharmacies? Please explain.

*[Probe, as needed, to understand what exactly the interviewee is proposing and what they think would improve if this change were implemented (e.g., do they think the change would improve pharmacy providers’ confidence in their ability to deliver PrEP; make PrEP delivery less burdensome for providers; increase privacy for clients?]*

Do you think **any pharmacy** could deliver PrEP or only pharmacies meeting **certain criteria**? Please explain.

*[If not covered in their response to the above:]*

- What kind of **training and support** do pharmacy providers need to be able to deliver PrEP well?

*[Probe, as needed]*

- Do you feel like the training you received on PrEP prepared you well to deliver it at your pharmacy?
  - What, if anything, would you change about the training you received on PrEP?
- Did you ever contact the study’s remote clinician?
  - *[If yes]* Did you find it helpful having a remote clinician available? Why or why not?
  - *[If no]* Is this because you never had any need to consult the remote clinician? *[Probe, as needed, to understand if they didn’t feel comfortable contacting the remote clinician, etc.]*
- Did you find that you needed any additional job aids or support—for example, from the research assistant or other members of the research study team—to deliver PrEP at the pharmacy? Please explain.
- What kind of **staffing** do you think pharmacies need to be able to deliver PrEP well? (How many per shift? What cadres?)
- What kind of **space** do you think pharmacies need to be able to deliver PrEP well?
- Are there other things you think a pharmacy needs in order to be able to deliver PrEP well?

**[ONLY FOR PROVIDERS WHO DID *NOT* PARTICIPATE IN THE PILOT EXTENSION]**

I’d like to discuss a medication called pre-exposure prophylaxis or “PrEP”, for short. Have you ever heard of PrEP before? *[Hand interviewee a printed copy of this description so they can follow along]:*

As you may know, in 2015, the Kenyan Ministry of Health approved the use of PrEP for HIV prevention. It is a daily oral pill. If taken consistently every day, PrEP is a safe and effective way for individuals not living with HIV to reduce their risk of acquiring HIV. Currently, PrEP is only being delivered in select healthcare facilities in Kenya.

The Ministry of Health is interested in potentially allowing PrEP to be delivered at private pharmacies, such as the one where you work.

To deliver PrEP, pharmacy providers would:

- Ask clients questions about their recent sexual behaviors to confirm they are at risk of getting HIV
- Ask clients if they have a history of certain medical conditions that might contraindicate PrEP safety
- Test clients for HIV to confirm they are HIV-negative
- Counsel clients about how to reduce their HIV risk and about PrEP, including potential side effects and the importance of adherence
- Dispense PrEP
- And, for clients returning for follow-up, ask questions about their adherence and any side effects

Imagine also that a remote clinician is available via phone to answer questions you might have while screening or managing PrEP clients.

If you were trained on how to deliver PrEP, would you be open to delivering PrEP at your pharmacy? Why or why not?

Do you think you would be able to deliver PrEP as described above? Why or why not?

*[Probe, if needed]* What, if anything, do you think would be challenging about delivering PrEP at your pharmacy?

**TOPIC 2: CAB-LA**

I’m now going to talk to you about a new form of PrEP called “long-acting cabotegravir” or “CAB-LA”. It is also sometimes called “injectable PrEP”.

Have you ever heard of “CAB-LA” or “injectable PrEP” before?

*[If yes]*

- Where did you hear about it?
- What have you heard about it?

Ok. I’m going to tell you a little bit about CAB-LA *[Hand interviewee a printed copy of this description so they can follow along]:*

**Description of long-acting cabotegravir (CAB-LA) or “injectable PrEP”**

- CAB-LA is a new form of PrEP that is not yet available in Kenya. It is currently being reviewed by the Pharmacy and Poisons Board.
- Like oral PrEP, CAB-LA is used to **prevent HIV** and must be taken **before** being exposed to HIV.
- Instead of taking one pill every day, people using CAB-LA get **one injection** **every two** months. It is given as an intramuscular injection in the **upper part of the client’s bum/buttocks**.
- The injection contains cabotegravir, an HIV integrase inhibitor.
- Like oral PrEP, CAB-LA is **more than 90% effective** at preventing HIV in the whole body, regardless of how one is exposed to HIV.
- Like oral PrEP, CAB-LA **does not protect against pregnancy or sexually transmitted infections**. So if a person using CAB-LA wants to protect against pregnancy and/or infections like chlamydia and gonorrhea, they would still need to use condoms and/or contraception.
- Before each CAB-LA injection, a client must be **tested for HIV** to confirm they are HIV-negative.
- Side effects from CAB-LA are usually limited to the injection site, which may be a little tender, bruised, or swollen for a couple of days after the injection.

I’d like you to imagine that the Pharmacy and Poisons Board has approved CAB-LA and is going to allow it to be delivered in pharmacies like the one where you work.

Please also imagine that clients interested in CAB-LA would still need to be screened for HIV risk, tested for HIV, and counseled.

Do you think it would be **feasible for pharmacies to deliver CAB-LA** injections? Why or why not?

*[If not covered in their response to the above:]*

- What would be some of the **challenges** to delivering CAB-LA in pharmacies?
  - Does your pharmacy offer any injections, like injectable contraception (Depo-Provera) or vaccinations? *[If yes]* Which ones?
    - *[If pharmacy does offer injections]* Do you give any of these injections yourself?
      - *[If yes]* How did you learn how to give injections? (Did you learn on-the-job? Did you take a course? Where? For how long?)
      - *[If no]* Who gives injections at the pharmacy? (What is their professional background, e.g., pharmacist, nurse, HTS counselor?)
  - Does your pharmacy currently have any way of disposing used sharps?
    - *[If yes]* How does your pharmacy dispose of used sharps currently?
    - *[If no]* How easy or hard do you think it would be for your pharmacy to dispose of used sharps?
  - If you were trained on how to give CAB-LA injections—which, again, go into the client’s upper buttocks regions—do you think you would be comfortable doing so at the pharmacy? Why or why not? *[If not, probe, as needed, to understand if discomfort is related to the injection site, to privacy concerns, etc.]*
  - How easy or hard do you think it would be to counsel clients about both CAB-LA and oral PrEP to inform their decision about which form of PrEP is right for them? *[Probe, as needed, to understand what they think would be easy or difficult].*
  - Do you think clients would be comfortable getting a CAB-LA injection at your pharmacy? Why or why not?
    - Is there anything a pharmacy could do to make clients more comfortable getting a CAB-LA injection there? Please explain.
- *[If not already answered]* What are some possible solutions to the challenges you’ve mentioned?

Earlier, you described the kinds of training and support you thought pharmacy providers need in order to deliver oral PrEP well. What, if any, **additional training or support** do you think pharmacy providers would need to deliver **CAB-LA** well?

*[Probe, as needed]*

- Are there any **core** **skills or competencies** that you think pharmacy providers would need additional training on in order to deliver CAB-LA?
- Are there any specific **job aids, tools, or other resources** that you think would help pharmacy providers deliver CAB-LA? Please explain.
- Do you think having a **remote clinician** available for consultation would be helpful for pharmacy providers delivering CAB-LA? Why or why not?

Do you think your pharmacy would need to be **staffed** differently to be able to deliver CAB-LA? *[If yes]* How so? *[Probe to understand if they think the pharmacy would need just more staff overall, more staff of particular cadre, more staff during certain times of day, etc.]*

Do you think the current **configuration of space** at your pharmacy would work for delivering CAB-LA or would you need to make some adjustments? Please explain.

Are there any additional **equipment, furniture, or other supplies** that you think your pharmacy would need in order to deliver CAB-LA?

Is there **anything else** you think a pharmacy needs in order to be able to deliver CAB-LA well?

If CAB-LA were available at pharmacies, what do you think would be the best way to **spread the word** about its availability there? *[Probe, as needed, to understand recommended platform, target audience, and/or key points to include in messaging.]*

How interested would you be in delivering CAB-LA at your pharmacy?

**TOPIC 3: 6-month PrEP Injection**

I’m going to tell you a little bit about another kind of PrEP injection not yet available. *[Hand interviewee a printed copy of this description so they can follow along]:*

**Description of 6-month PrEP injection:**

- A 6-month PrEP injection n is currently being investigated in Phase III clinical trials.
- It contains a drug called *lenacapavir,* which is the first of a new class of antiretrovirals—called capsid inhibitors—that disrupt HIV replication by interfering with the capsid shell of the HIV virus.
- Whereas CAB-LA injections are every two months (6 times per year), this 6-month PrEP injection is anticipated to be **every six months (so twice per year)**.
- This 6-month PrEP injection is subcutaneous and goes into the **abdomen**.

I’d like you to imagine that the clinical trials prove that this 6-month PrEP injection is as effective at preventing HIV as PrEP pills and CAB PrEP, and that the Pharmacy and Poisons Boards approves it for use in Kenya.

Do you think it would be **feasible for pharmacies to deliver** six-month PrEP injections? Why or why not?

*[If interviewee’s answer for the 6-month PrEP injection differs from what they said for CAB-LA, probe as needed to understand why they think the 6-month PrEP injection would be more/less feasible for pharmacies to deliver.]*

Whereas CAB-LA is an intramuscular injection that goes into the upper bum/buttocks, this 6-month PrEP injection is subcutaneous and goes into the abdomen. Do you think this would be easier for pharmacy providers to administer, harder for them to administer, or about the same? *[If easier or harder, probe to understand why and whether this has implications for the training/support pharmacy providers might need to administer the 6-month PrEP injection.]*

Clients who opt to get this 6-month PrEP injection would only need to return to the pharmacy twice per year. How easy or hard do you think it would be for clients to stick to this schedule? Do you have any suggestions as to how pharmacies could help these clients remember to return?

**TOPIC 4: Dapivirine vaginal ring**

I’m now going to talk to you about another new form of PrEP called the dapivirine vaginal ring or “PrEP ring”.

Have you ever heard of the dapivirine vaginal ring or “PrEP ring” before?

*[If yes]*

- Where did you hear about it?
- What have you heard about it?

Ok. I’m going to tell you a little bit about the PrEP ring *[Hand interviewee a printed copy of this description so they can follow along]:*

**Description of dapivirine vaginal ring or “PrEP ring”**

- The PrEP ring is a new form of PrEP for **women** that was recently approved by the Kenya Ministry of Health and is slowly being rolled out to public health facilities.
- Like oral PrEP, the ring is used to **prevent HIV** and must be worn **before** being exposed to HIV through **receptive vaginal sex**.
- The ring is made of flexible silicone rubber and contains *dapivirine*, a non-nucleoside reverse transcriptase inhibitor (NNRTI). The ring does not need to be stored in a refrigerator.
- A woman can insert the ring into her vagina by herself and leave it in for **28 days**. During these 28 days, the ring does not have to be taken out or cleaned, even after sex or menstruation. At the end of 28 days, she can take the ring out by herself and insert a new ring.
- The ring is about **50% effective at preventing HIV in the vagina**. It does not prevent HIV in other parts of the body, such as the anus.
- Like oral PrEP, the ring does not protect against pregnancy or sexually transmitted infections (STIs). So women using the ring who want to protect themselves against pregnancy and/or STIs, like chlamydia and gonorrhea, still need to use condoms and/or contraception.
- As with oral PrEP, women using the ring need to **get tested for HIV periodically** to ensure they are still HIV-negative.
- Side effects from the ring are uncommon. Some users report very mild side effects, like vaginal discomfort, that usually go away after a few days.

I’d like you to imagine that the Pharmacy and Poisons Board is going to allow the ring to be delivered in pharmacies like the one where you work.

Please also imagine that female clients interested in the ring would still need to be screened for HIV risk, tested for HIV, and counseled on how to insert the ring into the vagina; however, the insertion would not need to happen at the pharmacy. That is, the client could take the ring home to insert it herself.

Do you think it would be **feasible for pharmacies to deliver** the PrEP ring? Why or why not?

*[If not covered in their response to the above:]*

- What would be some of the **challenges** to delivering the PrEP ring in pharmacies?
  - Does your pharmacy sell the contraception ring (e.g., the Nuvaring)?
    - *[If yes]* Do you ever counsel clients on the contraception ring, including how to insert it?
      - *[If yes]* How comfortable do you feel doing this counseling?
      - *[If no]* Who does this counseling? (Are they male/female? What is their professional background, e.g., pharmacist, nurse, HTS counselor?)
  - If you were trained on how to deliver the PrEP ring, do you think you would be comfortable doing so at the pharmacy Why or why not? *[If not, probe, as needed, to understand if discomfort is related to privacy concerns, their own age or sex (especially if interviewee is male), etc.]*
  - How easy or hard do you think it would be to counsel clients about both oral PrEP and the PrEP ring to inform their decision about which form of PrEP is right for them?
  - Do you think female clients would be comfortable purchasing the PrEP ring at your pharmacy? Why or why not?
    - Is there anything a pharmacy could do to make female clients more comfortable purchasing the PrEP ring there? Please explain.
- *[If not already answered]* What are some possible solutions to the challenges you’ve mentioned?

Are there any specific **job aids, tools, or other resources** that you think would help pharmacy providers deliver the PrEP ring? Please explain.

If the PrEP ring were available at pharmacies, what do you think would be the best way to **spread the word** about its availability there? *[Probe, as needed, to understand recommended platform, target audience, and/or key points to include in messaging.]*

How interested would you be in delivering the PrEP ring at your pharmacy?

**TOPIC 4: Closing thoughts**

Is there anything else you’d like to share about your thoughts or experience delivering oral PrEP at the pharmacy or ways you think pharmacies might succeed at delivering CAB-LA and the dapivirine vaginal ring?

We’ve covered all the topics I wanted to discuss today. Thanks very much for your time. I’m going to shut off the recorder now.

If you have any additional questions about oral PrEP, CAB-LA or the PrEP ring, I’d be happy to answer those or tell you where you can find more information.

**[Mark interview end time on page 1 (Item I).]**

**In-Depth Interview Guide: Stakeholders**

**Interviewer instructions: Administer informed consent. Once signed, begin guide.**

**0.0 Interview Information**

***Fill out items A through I prior to starting interview.***

(q) **Informed consent has been administered**: YES	/	NO

*If consent form has not been signed by participant,*

*interview must not proceed.*

(r) **Interview ID**: _____________________________

*Format: Interviewee Type-DDMMYY-Number interview conducted that day*

*where “S” = Stakeholder (e.g., “S-150423-02” represents the second stakeholder interviewed on 15 Apr 2023)*

(s) **Date of interview**: ____/_____/_______

*Format: DD/MM/YYYY*

(t) **Location of interview**: __________________________________________

(u) **Interviewer’s full name**: _________________________________________

(v) **Interview start time**: ____________________

*Format: HH:MM am or pm*

(w) **Interview end time**: _____________________

*Format: HH:MM am or pm*

**Facilitator introduction: [DO NOT READ; GUIDE ONLY]**

Hello. My name is _______, and I am a ________, working at _______. Thank you for taking the time to talk with me today.

The purpose of this interview is to understand your thoughts about new, long-acting forms of PrEP and the idea of potentially delivering these via private pharmacies in Kenya.

There are no right or wrong answers to these questions. People have different views and we are interested to learn more about these experiences from you. Today, you are in the role of a teacher and I am here to learn from you since you are an expert in your own life experiences and opinions.

This interview should take around an hour to complete. Please let me know if at any time you have questions, if something I say is not clear, or if you need to take a break.

| 23. **[*INTERVIEWER: Mark the participant’s sex.*]** | *☐ Male*  *☐ Female* |
| --- | --- |
| ***START INTERVIEW HERE.*** | |
| I’d like to start off by asking you some basic questions about yourself. | |
| 24. How old are you? | _________ years old |
| 25. What is the name of the organization you work for? | ___________________________ |
| 26. Which of the following best categorizes your organization? | *☐ Policymaker*  *☐ Regulator*  *☐ Advocacy group*  *☐ Implementing partner*  *☐ Funder*  *☐ Other: _____________________________* |
| 27. How many years have you worked for this organization? | ___________ years |
| 28. What is your current role (job title) at this organization? | ______________________________ |

**TOPIC 1: PrEP policy & decision-making in Kenya**

I will now start the audio recording.

Part of the reason why we were interested in interviewing you is because your organization plays a key role in PrEP policy and decision-making in Kenya.

Can you describe what your organization sees as its **key mission or priorities related to PrEP** in Kenya?

*[Probe, if needed]* What kind of activities related to PrEP does your organization carry out?

*[Only if interviewee is from MOH or PPB]* Can you tell me a bit about how your organization makes decisions related to PrEP?

*[Probes, if needed]*

- Are there specific kinds of evidence your organization likes to see before changing policy? Please explain.
- What influence, if any, do organizations *outside of Kenya*—like the ministries of health of other countries or the World Health Organization—have on how your organization makes decisions related to PrEP? *[Probe, as needed, to understand which organizations they think influence PrEP policymaking in Kenya and how.]*

What **concerns**, if any, does your organization have about PrEP?

*[Probe, as needed, to understand if the concerns are related to PrEP access/equity, safety, cost/cost-effectiveness, impact on HIV incidence, etc.]*

What other organizations or individuals do you think play a key role in PrEP policy and decision-making in Kenya?

*[For each organization/individual mentioned:]* What makes you say that [Organization X/Individual X] plays a key role in PrEP policy and decision-making in Kenya? (What do they do? How are their actions or inactions consequential for how PrEP is implemented in Kenya?)

**TOPIC 2: Dapivirine vaginal ring**

As I’m sure you’re aware, several new forms of long-acting PrEP have come out in recent years. I’d like to ask you a bit about a couple of them.

Let’s start with the dapivirine vaginal ring.

Have you heard of the dapivirine vaginal ring?

*[If yes]* What have you heard about it?

*[Regardless of whether interviewee has heard of the ring before or not:] Ok. Just to make sure that we’re all on the same page, let me share with you a brief description of the dapivirine vaginal ring. You can read through it yourself. [Hand interviewee a printed copy of the description and let them read through it on their own.]*

**Description of dapivirine vaginal ring or “PrEP ring”**

- The PrEP ring is a new form of PrEP for **women** that was recently approved by the Kenya Ministry of Health and is slowly being rolled out to public health facilities.
- Like oral PrEP, the ring is used to **prevent HIV** and must be worn **before** being exposed to HIV through **receptive vaginal sex**.
- The ring is made of flexible silicone rubber and contains *dapivirine*, a non-nucleoside reverse transcriptase inhibitor (NNRTI). The ring does not need to be stored in a refrigerator.
- A woman can insert the ring into her vagina by herself and leave it in for **28 days**. During these 28 days, the ring does not have to be taken out or cleaned, even after sex or menstruation. At the end of 28 days, she can take the ring out by herself and insert a new ring.
- The ring is about **50% effective at preventing HIV in the vagina**. It does not prevent HIV in other parts of the body, such as the anus.
- Like oral PrEP, the ring does not protect against pregnancy or sexually transmitted infections (STIs). So women using the ring who want to protect themselves against pregnancy and/or STIs, like chlamydia and gonorrhea, still need to use condoms and/or contraception.
- As with oral PrEP, women using the ring need to **get tested for HIV periodically** to ensure they are still HIV-negative.
- Side effects from the ring are uncommon. Some users report very mild side effects, like vaginal discomfort, that usually go away after a few days.

In general, do you think the dapivirine ring is a good HIV prevention option to make available to women in Kenya? Why or why not?

*[Probe, as needed, to understand any conditions they have [e.g., if they think it’s only good for certain populations of women], any concerns they have about the ring, and/or additional information they wish they knew about the ring.]*

Do your think your organization would be in favor of allowing **private pharmacies** to deliver the dapivirine ring? Why or why not?

*[If not covered in response to the above]*

- *[If interviewee thinks organization would be in favor]*
  - What benefits do you think would come from making the ring available at private pharmacies?
  - What support or resources do you think your organization could commit to making the ring available at private pharmacies.
- *[If interviewee thinks organization would not be in favor]*
  - What concerns would your organization have about making the ring available at private pharmacies?
- *[Only for interviewees from MOH or PPB]* What kinds of **policy changes**, if any, would be needed to allow private pharmacies to deliver the ring? *[If needed, probe: “If private pharmacies had access to dapivirine ring stock, could they legally deliver it right now or would the MOH and/or PPB need to change some rules to allow this?”]*

*[For each policy change mentioned:]*

- Who has the authority to change policy on [X, e.g., which cadres are allowed to prescribe PrEP]? *[Probe, as needed, to understand which organizations/individuals have the authority to make the policy changes mentioned].*
- How likely do you think [policymaking organization, e.g., MOH/PPB] is to change policy on [X]? *[Probe, as needed, to understand why they think the policy change is likely or unlikely and under what circumstances, if any, they think the policy change might go through.]*
  - *[If interviewee thinks this policy change is likely]* And do you think [policymaking organization, e.g., MOH/PPB] might change policy on [X] anytime soon? What is a realistic timeframe for this change? *[Probe to understand if they think it’s within the next year, the next 2-5 years, etc.]*

Do you foresee any other **challenges** to having private pharmacies deliver the dapivirine ring? Please explain. *[If needed, probe to understand if the challenges they anticipate are related to privacy at the pharmacy (physical space/layout) and client comfort, cost for clients, supply chain/stock-outs, pharmacy providers’ ability to counsel clients about the ring, etc.]*

Can you think of **any potential solutions** to the challenges you’ve raised? *[Probe about the specific challenges the interviewee raised. For example, if the interviewee said that most pharmacy providers don’t know about the ring, ask them if they think they could be trained on this and, if so, what they think that training should entail (content, duration, job aids, etc.) Or if the interviewee said that cost may be an issue, ask them whether they think some clients might be willing to pay and their thoughts on the MOH, PEPFAR, or other organizations potentially subsidizing the cost for clients to get the ring at private pharmacies.)*

*[If not covered in their responses to previous questions]:* Do you think **any pharmacy** could deliver the dapivirine ring or only pharmacies meeting **certain criteria**? Please explain.

**TOPIC 3: CAB-LA**

I’m now going to talk to you about a new form of PrEP called “long-acting cabotegravir” or “CAB-LA”. It is also sometimes called “injectable PrEP”.

Have you ever heard of “CAB-LA” or “injectable PrEP” before?

*[If yes]* What have you heard about it?

*[Regardless of whether interviewee has heard of CAB-LA before or not:] Ok. Just to make sure that we’re all on the same page, let me share with you a brief description of CAB-LA. You can read through it yourself. [Hand interviewee a printed copy of the description and let them read through it on their own.]*

**Description of long-acting cabotegravir (CAB-LA) or “injectable PrEP”**

- CAB-LA is a new form of PrEP that is not yet available in Kenya. It is currently being reviewed by the Pharmacy and Poisons Board.
- Like oral PrEP, CAB-LA is used to **prevent HIV** and must be taken **before** being exposed to HIV.
- Instead of taking one pill every day, people using CAB-LA get **one injection** **every two** months. It is given as an intramuscular injection in the **upper part of the client’s bum/buttocks**.
- The injection contains cabotegravir, an HIV integrase inhibitor.
- Like oral PrEP, CAB-LA is **more than 90% effective** at preventing HIV in the whole body, regardless of how one is exposed to HIV.
- Like oral PrEP, CAB-LA **does not protect against pregnancy or sexually transmitted infections**. So if a person using CAB-LA wants to protect against pregnancy and/or infections like chlamydia and gonorrhea, they would still need to use condoms and/or contraception.
- Before each CAB-LA injection, a client must be **tested for HIV** to confirm they are HIV-negative.
- Side effects from CAB-LA are usually limited to the injection site, which may be a little tender, bruised, or swollen for a couple of days after the injection.

In general, do you think CAB-LA is a good HIV prevention option to make available in Kenya? Why or why not?

*[Probe, as needed, to understand any conditions they have [e.g., if they think it’s only good for certain populations], any concerns they have about CAB-LA, and/or additional information they wish they knew about CAB-LA.]*

If CAB-LA gets approved for use in Kenya, do you think your organization would be in favor of allowing **private pharmacies** to deliver CAB-LA? Why or why not?

*[If not covered in response to the above]*

- *[If interviewee thinks organization would be in favor]*
  - What benefits do you think would come from making CAB-LA available at private pharmacies?
  - What support or resources do you think your organization could commit to making CAB-LA available at private pharmacies.
- *[If interviewee thinks organization would not be in favor]*
  - What concerns would your organization have about making CAB-LA available at private pharmacies?
- *[Only for interviewees from MOH or PPB]* What kinds of **policy changes**, if any, would be needed to allow private pharmacies to deliver CAB-LA? *[If needed, probe: “If private pharmacies had access to CAB-LA stock, could they legally deliver it right now or would the MOH and/or PPB need to change some rules to allow this?”]*

*[For each policy change mentioned:]*

- Who has the authority to change policy on [X, e.g., which cadres are allowed to deliver injections]? *[Probe, as needed, to understand which organizations/individuals have the authority to make the policy changes mentioned].*
- How likely do you think [policymaking organization, e.g., MOH/PPB] is to change policy on [X]? *[Probe, as needed, to understand why they think the policy change is likely or unlikely and under what circumstances, if any, they think the policy change might go through.]*
  - *[If interviewee thinks this policy change is likely]* And do you think [policymaking organization, e.g., MOH/PPB] might change policy on [X] anytime soon? What is a realistic timeframe for this change? *[Probe to understand if they think it’s within the next year, the next 2-5 years, etc.]*

Do you foresee any other **challenges** to having private pharmacies deliver CAB-LA? Please explain. *[If needed, probe to understand if the challenges they anticipate are related to privacy at the pharmacy (physical space/layout) and client comfort, cost for clients, supply chain/stock-outs, pharmacy providers’ ability to administer injections, etc.]*

Can you think of **any potential solutions** to the challenges you’ve raised? *[Probe about the specific challenges the interviewee raised. For example, if the interviewee said that most pharmacy providers don’t know about CAB-LA, ask them if they think they could be trained on this and, if so, what they think that training should entail (content, duration, job aids, etc.) Or if the interviewee said that cost may be an issue, ask them whether they think some clients might be willing to pay and their thoughts on the MOH, PEPFAR, or other organizations potentially subsidizing the cost for clients to get CAB-LA at private pharmacies.)*

*[If not covered in their responses to previous questions]:* Do you think **any pharmacy** could deliver CAB-LA or only pharmacies meeting **certain criteria**? Please explain.

**TOPIC 4: 6-month PrEP Injection**

I’m going to tell you a little bit about another kind of PrEP injection not yet available. *[Hand interviewee a printed copy of this description so they can follow along]:*

**Description of 6-month PrEP injection:**

- A 6-month PrEP injection is currently being investigated in Phase III clinical trials.
- It contains a drug called *lenacapavir,* which is the first of a new class of antiretrovirals—called capsid inhibitors—that disrupt HIV replication by interfering with the capsid shell of the HIV virus.
- Whereas CAB-LA injections are every two months (6 times per year), this 6-month PrEP injection is anticipated to be **every six months (so twice per year)**.
- This 6-month PrEP injection is subcutaneous and goes into the **abdomen**.

I’d like you to imagine that the clinical trials prove that this 6-month PrEP injection is as effective at preventing HIV as oral PrEP and CAB-LA, and that the Pharmacy and Poisons Boards approves it for use in Kenya.

What are your thoughts on private pharmacies **delivering 6-month PrEP injections**?

*[If interviewee’s answer for the 6-month PrEP injection differs from what they said for CAB-LA, probe as needed to understand why. For example, why do they have more concerns/fewer concerns about pharmacies delivering 6-month PrEP injections or why they think it would think more feasible or less feasible for pharmacies to deliver the six-month PrEP injection, etc.]*

**TOPIC 5: Recommendations for our pilot study**

As you may know, over the past 2 years, we at [PHRD/KEMRI] conducted a pilot study of pharmacy-delivered oral PrEP. We trained pharmacists and pharmaceutical technologists to use a standardized checklist to screen clients for HIV risk behaviors; conduct a medical safety assessment to ensure they didn’t have any contraindications to PrEP; assist clients with HIV testing; and dispense PrEP. The first phase of the study ran for 1 year—from November 2020 to October 2021—at 4 pharmacies in Kiambu and Kisumu Counties. Those 4 pharmacies initiated about 300 clients on PrEP, and 41% of them refilled PrEP at the pharmacy at least once during the study. Then from February to July of 2022, we ran an extension of this study at 12 pharmacies in the same two counties. Over that 6-month period, the 12 pharmacies initiated about 850 clients on PrEP and 200 on post-exposure prophylaxis (PEP). And 72% of PrEP clients refilled PrEP at the pharmacy at least once.

We’re now preparing to conduct an 18-month pilot study, following approvals from the Scientific Ethics Review Unit, the Ministry of Health, and the Pharmacy and Poisons Board, among others. Our plan is that, for the first 6 months of this study, trained pharmacy providers will deliver both oral PrEP and the dapivirine ring. Then we plan to introduce injectable PrEP for the last 12 months of the study. (We will discuss injectable PrEP in more detail in just a few minutes). Regardless of the type of PrEP they choose, participants will not pay anything to receive PrEP at the pharmacy. ***[Hand interviewee the one-page description of the CAB-LA pilot study]***

Is there anything in particular that you and your organization would be **interested to learn** from this study? Please explain.

*[Probe, as needed, to understand if they’re interested in which PrEP form clients choose, whether they switch products during the study, what demographic is reached, rates of seroconversion, etc.]*

Do you have any **recommendations for our study** or **things you think we should keep in mind as we prepare for this study**?

*[Probes, if needed]*

- For example, do you have any recommendations for how we should **prepare pharmacy providers** or how we should **support them** to deliver these 3 kinds of PrEP?
- Do you have any recommendations for **how to engage potential clients** in pharmacy-based PrEP services?

**TOPIC 6: Closing thoughts**

Is there anything else you’d like to share about your thoughts on private pharmacies delivering oral PrEP, the dapivirine ring, and/or CAB-LA?

We’ve covered all the topics I wanted to discuss today. Thanks very much for your time. I’m going to shut off the recorder now.

If you have any additional questions about the dapivirine ring or CAB-LA, I’d be happy to answer those or tell you where you can find more information.

**[Mark interview end time on page 1 (Item G).]**
